# Integrated immune networks in SARS-CoV-2 infected pregnant women reveal differential NK cell and unconventional T cell activation

**DOI:** 10.1101/2021.08.21.21262399

**Authors:** Jennifer R Habel, Brendon Y Chua, Lukasz Kedzierski, Kevin J Selva, Timon Damelang, Ebene R Haycroft, Thi HO Nguyen, Hui-Fern Koay, Suellen Nicholson, Hayley McQuilten, Xiaoxiao Jia, Lilith F Allen, Luca Hensen, Wuji Zhang, Carolien E van de Sandt, Jessica A Neil, Fatima Amanat, Florian Krammer, Kathleen Wragg, Jennifer A Juno, Adam K Wheatley, Hyon-Xhi Tan, Gabrielle Pell, Jennifer Audsley, Irani Thevarajan, Justin Denholm, Kanta Subbarao, Dale I Godfrey, Allen C Cheng, Steven YC Tong, Katherine Bond, Deborah A Williamson, Fiona James, Natasha E Holmes, Olivia C Smibert, Jason A Trubiano, Claire L Gordon, Amy W Chung, Clare L Whitehead, Stephen J Kent, Martha Lappas, Louise C Rowntree, Katherine Kedzierska

**Affiliations:** Department of Microbiology and Immunology, University of Melbourne, at the Peter Doherty Institute for Infection and Immunity, Melbourne, Victoria 3000, Australia; Global Station for Zoonosis Control, Global Institution for Collaborative Research and Education (GI-CoRE), Hokkaido University, Sapporo, Japan; Faculty of Veterinary and Agricultural Sciences, University of Melbourne, Melbourne, Victoria 3000, Australia; Victorian Infectious Diseases Reference Laboratory, The Royal Melbourne Hospital at The Peter Doherty Institute for Infection and Immunity, Melbourne, Victoria 3000, Australia; Department of Microbiology, Icahn School of Medicine at Mount Sinai, New York, NY, USA; Graduate School of Biomedical Sciences, Icahn School of Medicine at Mount Sinai, New York, NY, USA; ARC Centre of Excellence in Convergent Bio-Nano Science and Technology, University of Melbourne, Melbourne, Victoria 3000, Australia; Mercy Perinatal Research Centre, Mercy Hospital for Women, Heidelberg, Victoria 3084, Australia; Department of Infectious Diseases, University of Melbourne at the Peter Doherty Institute for Infection and Immunity, Melbourne, Victoria 3000, Australia; Victorian Infectious Diseases Service, The Royal Melbourne Hospital at the Peter Doherty Institute for Infection and Immunity, Melbourne, Victoria 3000, Australia; World Health Organisation (WHO) Collaborating Centre for Reference and Research on Influenza, at The Peter Doherty Institute for Infection and Immunity, Melbourne, Victoria 3000, Australia; School of Public Health and Preventive Medicine, Monash University, Melbourne, Victoria, Australia; Infection Prevention and Healthcare Epidemiology Unit, Alfred Health, Melbourne, Victoria, Australia; Menzies School of Health Research and Charles Darwin University, Darwin, Northern Territory, Australia; Department of Microbiology, Royal Melbourne Hospital, at The Peter Doherty Institute for Infection and Immunity, Melbourne, Victoria 3000, Australia; Department of Infectious Diseases, Austin Health, Heidelberg, Victoria 3084, Australia; Department of Critical Care, University of Melbourne, Parkville, Victoria 3000, Australia; Data Analytics Research and Evaluation (DARE) Centre, Austin Health and University of Melbourne, Heidelberg, Victoria 3084, Australia; Centre for Antibiotic Allergy and Research, Department of Infectious Diseases, Austin Health, Heidelberg, Victoria 3084, Australia; Department of Infectious Diseases, Peter McCallum Cancer Centre, Melbourne, Victoria 3000, Australia; National Centre for Infections in Cancer, Peter McCallum Cancer Centre, Melbourne, Victoria 3000, Australia; Department of Medicine (Austin Health), University of Melbourne, Heidelberg, Victoria 3084, Australia; Department of Obstetrics and Gynaecology, University of Melbourne, Parkville, Victoria 3052, Australia; Pregnancy Research Centre, The Royal Women’s Hospital, Parkville, Victoria, Australia; Melbourne Sexual Health Centre, Infectious Diseases Department, Alfred Health, Central Clinical School, Monash University, Melbourne, Victoria, Australia; Obstetrics, Nutrition and Endocrinology Group, Department of Obstetrics and Gynaecology, University of Melbourne, Victoria, Australia

## Abstract

Although pregnancy poses a greater risk for severe COVID-19, the underlying immunological changes associated with SARS-CoV-2 infection during pregnancy are poorly understood. We defined immune responses to SARS-CoV-2 in pregnant and non-pregnant women during acute and convalescent COVID-19 up to 258 days post symptom onset, quantifying 217 immunological parameters. Additionally, matched maternal and cord blood were collected from COVID-19 convalescent pregnancies. Although serological responses to SARS-CoV-2 were similar in pregnant and non-pregnant women, cellular immune analyses revealed marked differences in key NK cell and unconventional T cell responses during COVID-19 in pregnant women. While NK cells, γδ T cells and MAIT cells displayed pre-activated phenotypes in healthy pregnant women when compared to non-pregnant age-matched women, activation profiles of these pre-activated NK and unconventional T cells remained unchanged at acute and convalescent COVID-19 in pregnancy. Conversely, activation dynamics of NK and unconventional T cells were prototypical in non-pregnant women in COVID-19. In contrast, activation of αβ CD4^+^ and CD8^+^ T cells, T follicular helper cells and antibody-secreting cells was similar in pregnant and non-pregnant women with COVID-19. Elevated levels of IL-1β, IFN-γ, IL-8, IL-18 and IL-33 were also found in pregnant women in their healthy state, and these cytokine levels remained elevated during acute and convalescent COVID-19. Collectively, our study provides the first comprehensive map of longitudinal immunological responses to SARS-CoV-2 infection in pregnant women, providing insights into patient management and education during COVID-19 pregnancy.

## INTRODUCTION

Severe acute respiratory syndrome coronavirus 2 (SARS-CoV-2) emerged in late 2019, causing a pandemic that has resulted in hundreds of million infections and >4 million deaths globally (Dong et al., 2020). As COVID-19 case numbers continue to rise, understanding immune responses to SARS-CoV-2, especially in high-risk groups, is of critical importance to guide treatment and vaccine strategies. The majority of immunological studies on COVID-19 have largely focused on the correlates of disease severity in previously healthy individuals across different age groups (Juno et al., 2020c; Koutsakos et al., 2021; Thevarajan et al., 2020). Most COVID-19 patients develop prototypical broad, robust and transient anti-viral immune responses to SARS-CoV-2 infection (Koutsakos et al., 2021; Kuri-Cervantes et al., 2020; Laing et al., 2020; Long et al., 2020; Mathew et al., 2020; Thevarajan et al., 2020), with abundant SARS-CoV-2-specific antibodies, B cell and T cell responses (Amanat et al., 2020; Grifoni et al., 2020; Habel et al., 2020; Juno et al., 2020b; Weiskopf et al., 2020), establishing long-lasting memory (Nguyen et al., 2014; Nguyen et al., 2021; Rodda et al., 2021; Rowntree et al., 2021; Wheatley et al., 2021). Hyperactivation of innate and adaptive immune responses as well as blood hypercytokinemia are characteristic of severe disease (Koutsakos et al., 2021; Kuri-Cervantes et al., 2020; Lucas et al., 2020; Mathew et al., 2020). There is, however, still a paucity of data on immune responses to SARS-CoV-2 infection in groups vulnerable to poor outcomes following infection, especially pregnant women.

Pregnant women are considered to be a vulnerable group for SARS-CoV-2 infection due to physiological and immunological changes occurring during gestation (Zambrano et al., 2020). Studies to date associate COVID-19 during pregnancy with an increased risk of intensive care unit (ICU) admission, invasive ventilation and extracorporeal membrane oxygenation (ECMO) compared to non-pregnant women of reproductive age (Allotey et al., 2020; Zambrano et al., 2020). In comparison to non-pregnant women with COVID-19, pregnant women with COVID-19 are at an increased risk of death, sepsis, mechanical ventilation, ICU admission, shock, acute renal failure and thromboembolic disease (Ko et al., 2021). Additionally, COVID-19 during pregnancy has been linked to an increased risk of pre-eclampsia and gestational hypertension, resulting in a greater risk of adverse pregnancy outcomes (Papageorghiou et al., 2021). Nonetheless, others have shown that pregnant women commonly have mild or asymptomatic SARS-CoV-2 infection (Crovetto et al., 2020). Pregnancy presents unique physiological and immunological states which are required to maintain a viable and healthy fetus, while still protecting the mother from infections. Gestational immune alterations can impair anti-viral responses, leading to severe disease such as that observed in the 1918, 1957 and 2009 influenza pandemics (Creanga et al., 2010; Eickhoff et al., 1961; Harris, 1919; Louie et al., 2010; Siston et al., 2010).

To date, published evidence shows that pregnant women who had COVID-19 produced SARS-CoV-2-specific antibodies, of which SARS-CoV-2-specific IgG antibodies were transferred to the cord blood (Atyeo et al., 2021; Edlow et al., 2020; Jang et al., 2021; Wang et al., 2021). A systematic review of clinical laboratory findings determined that a low white blood cell count was the only significant difference between the pregnant and non-pregnant COVID-19 immune responses (Areia and Mota-Pinto, 2020). However, pregnant or post-partum women who had recovered from COVID-19 had lower T follicular helper type 17 cells (T_FH_17), memory B cells, total and ‘virus’-specific (CD56^+^NKP46^+^) NK cells compared to healthy pregnant women (Zhao et al., 2021). Cytokine profiles differed between healthy pregnant women and those with COVID-19 (Chen et al., 2021; Zhao et al., 2021). Despite reports to date on specific immune parameters, a comprehensive analysis of immune perturbations in early and late stages of COVID-19 during pregnancy is lacking.

Our present study fills this knowledge gap and investigates the breadth of innate, adaptive and humoral immune responses to SARS-CoV-2 infection in pregnant women. Additionally, cord blood from convalescent COVID-19 pregnancies was assessed for SARS-CoV-2-specific antibodies, and placenta cellular compartments were examined for differential immune cell activation in COVID-19 and healthy pregnancies. We recruited 101 women to define immune responses to SARS-CoV-2 in pregnant and non-pregnant women during acute and convalescent COVID-19 up to 258 days post-disease onset, quantifying 217 immunological parameters. We provide the first comprehensive map of longitudinal immunological responses in COVID-19 pregnant women during acute and convalescent phases of SARS-CoV-2 infection. Our longitudinal comparisons revealed specifically lack of γδ T cell, MAIT and NK cell activation in pregnant women during acute COVID-19, as a result of their pre-activated profile during the healthy state. In contrast, activation of classical αβ CD4^+^ and CD8^+^ T cells, T follicular helper cells (T_FH_), antibody-secreting cells (ASC) and SARS-CoV-2-specific antibody patterns were similar across the groups. Differences in IL-1β, IFN-γ, IL-8, IL-18 and IL-33 levels were evident in a healthy state in pregnancy, and these cytokines remained elevated during acute and convalescent COVID-19. Taken together, our comprehensive analysis of immune dysfunction following COVID-19 in pregnancy provides key insights which can potentially inform patient management and education during COVID-19 in pregnancy.

## RESULTS

### COVID-19 pregnancy cohort demographics and study design

We recruited a total of 101 women into our study to understand cellular and humoral immune responses to SARS-CoV-2 during pregnancy (Fig 1A, Supp Table 1 and 2). Blood samples were collected from 19 pregnant women with PCR-confirmed COVID-19 during their pregnancy; 8 pregnant women with acute COVID-19 (<21 days post symptom onset) and 13 pregnant women at convalescence (≥21 days post symptom onset). As controls, blood samples were collected from 21 healthy pregnant women with no history of COVID-19. To define any alterations in the immune response to COVID-19 during pregnancy, we recruited 25 non-pregnant women with acute or convalescent PCR-confirmed COVID-19, and 37 healthy non-pregnant women (Fig 1A). Similar proportions of pregnant and non-pregnant women were located at home (50.0% vs 61.5%, respectively), in the hospital ward (45.0% vs 30.8%, respectively) or in the intensive care unit (ICU; 5.0% vs 7.7%, respectively) (Fig 1B). Pregnant women with acute or convalescent COVID-19 were recruited between 1-258 days post symptom onset, with 47% (n=9) and 53% (n=10) being in their second or third trimester, respectively (Fig 1C). Non-pregnant women with COVID-19 were recruited between 2-205 days post symptom onset (Fig 1D). There were no significant differences in the days post symptom onset at sample collection between acute pregnant and non-pregnant groups (median of 8 and 8.5 days, respectively) and convalescent pregnant and non-pregnant groups (median of 107 and 86 days, respectively) (Fig 1E). The ages of pregnant and non-pregnant women in the healthy, acute or convalescent groups were similar, ranging from 20-49 years (Fig 1F). The average gestation of pregnant women with COVID-19 was comparable to healthy pregnant women (Fig 1G).

**Figure 1.**
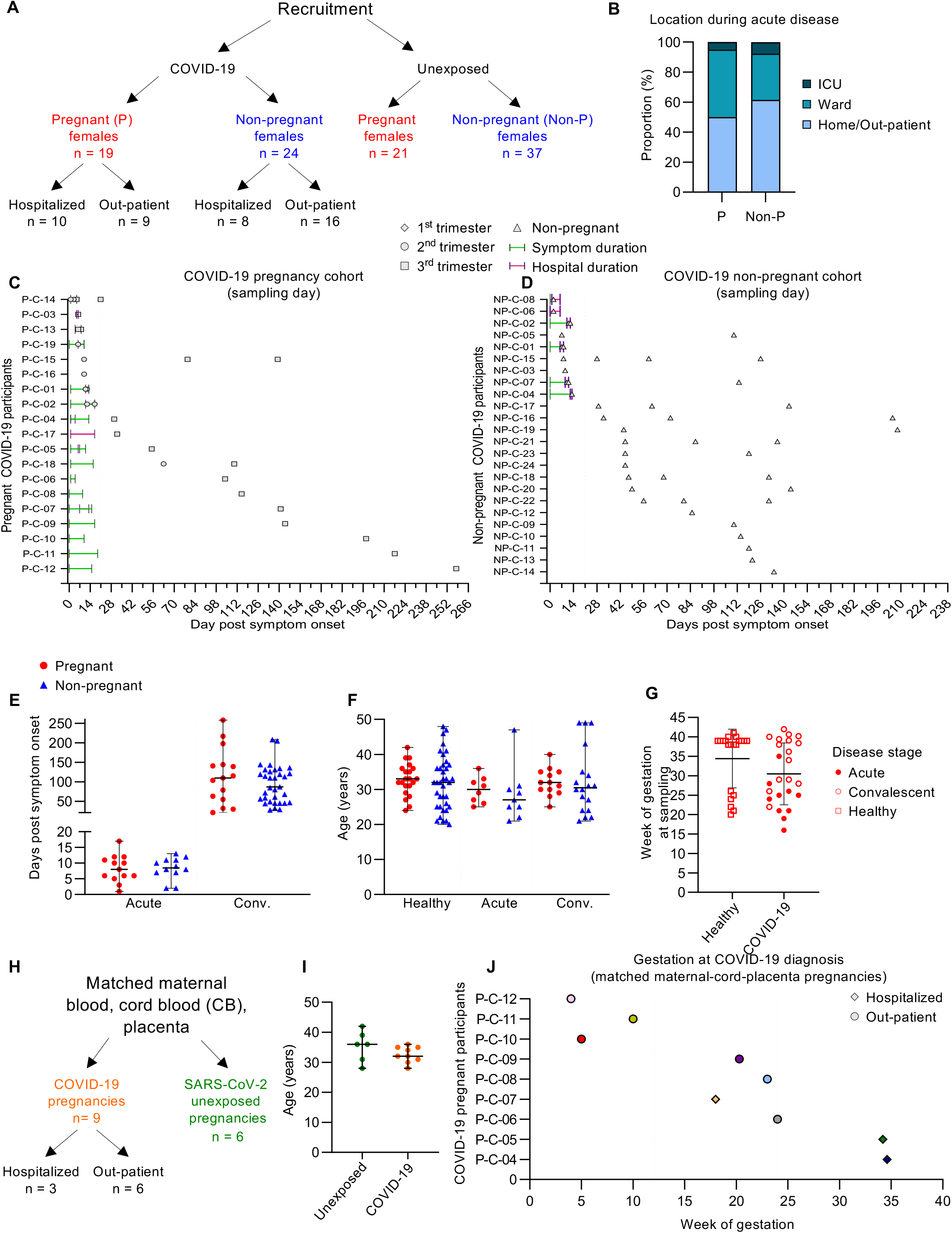
COVID-19 pregnancy cohort. (A) Schematic depicting recruitment of pregnant and non-pregnant women with acute or convalescent COVID-19 or who were not exposed to SARS-CoV-2. (B) Location during acute COVID-19 for pregnant (P) and non-pregnant (Non-P) donors. (C-D) Timeline showing days post disease symptom onset at which blood samples were collected for (C) pregnant and (D) non-pregnant women who had COVID-19. (E) Median days post symptom onset in acute (P n=13, Non-P n=12) and convalescent (P n=15, Non-P n=33) pregnant and non-pregnant donors. Donors with longitudinal sampling are represented for each timepoint collected. (F) Median age of pregnant and non-pregnant healthy (n=21 and 37), acute (n=8 and 9) or convalescent (n=13 and 18) COVID-19 donors. Median and range are shown. (G) Gestational age in weeks for healthy (n=21) and COVID-19 (n=25) pregnant donors. Means and standard deviations are shown. (H) Schematics showing a sub-group of recovered COVID-19 (n=9) or healthy (n=6) pregnant donors from whom matched cord blood and placenta tissue were collected in addition to maternal blood. Median age in years for healthy (n=6) and COVID-19 pregnant donors (n=9). Median and range are shown. (J) Timeline showing the week of gestation at which each pregnant donor became symptomatic with COVID-19 (n=9).

Matched maternal blood, cord blood and placenta tissue were collected at the birth time-point in a subset of COVID-19 (n=9) and SARS-CoV-2 unexposed (n=6) pregnancies (Supp Table 2). Of the 9 COVID-19 pregnancies, 3 were admitted to hospital and 6 were at home during their acute disease (Fig 1H). The ages of women with COVID-19 or unexposed pregnancy were not statistically different (Fig 1I). The gestational age at which COVID-19 was diagnosed ranged from 4-34 weeks, with 33.3% (n=3), 44.4% (n=4) and 22.2% (n=2) in the first, second or third trimester, respectively (Fig 1J). Additionally, 12 non-matched cord blood samples from healthy pregnancies formed a control group.

### Comparable RBD-specific and neutralizing antibodies in pregnant and non-pregnant women

As SARS-CoV-2 antibodies are associated with protection from repeated infection (Harvey et al., 2021), we assessed humoral responses to SARS-CoV-2 in pregnant and non-pregnant women through the detection of RBD-specific IgG, IgM and IgA antibodies by ELISA (Amanat et al., 2020; Koutsakos et al., 2021; Rowntree et al., 2021) (Fig 2A-F), while the surrogate virus neutralization assay (sVNT) was used to measure neutralizing activity of SARS-CoV-2-specific antibodies in COVID-19 patients (Nicholson et al., 2021; Rowntree et al., 2021; Tan et al., 2020) (Fig 2G). Overall, we detected no differences in the titres of RBD-specific IgG, IgM or IgA between pregnant and non-pregnant women with acute or convalescent COVID-19 (Fig 2B). Similarly, no significant differences in the avidity of RBD-IgG and RBD-IgM antibodies were found between pregnant and non-pregnant women when a urea-mediated dissociation ELISA was performed for donors who were bled sequentially (Fig 2C). Similar proportions of pregnant and non-pregnant donors seroconverted for IgG, IgM, IgA (Fig 2D), with the kinetics of RBD-IgG-specific titres also greatly overlapping (Fig 2E). Importantly, there were no differences in the proportion of ACE2-RBD inhibition detected by sVNT between pregnant and non-pregnant women with acute or convalescent COVID-19 (Fig 2F). The similarity in antibody titres between pregnant and non-pregnant women demonstrates that the production of SARS-CoV-2-specific antibodies is not impaired during pregnancy, and importantly suggests that women who had COVID-19 during pregnancy generate humoral protection from future re-infections.

**Figure 2.**
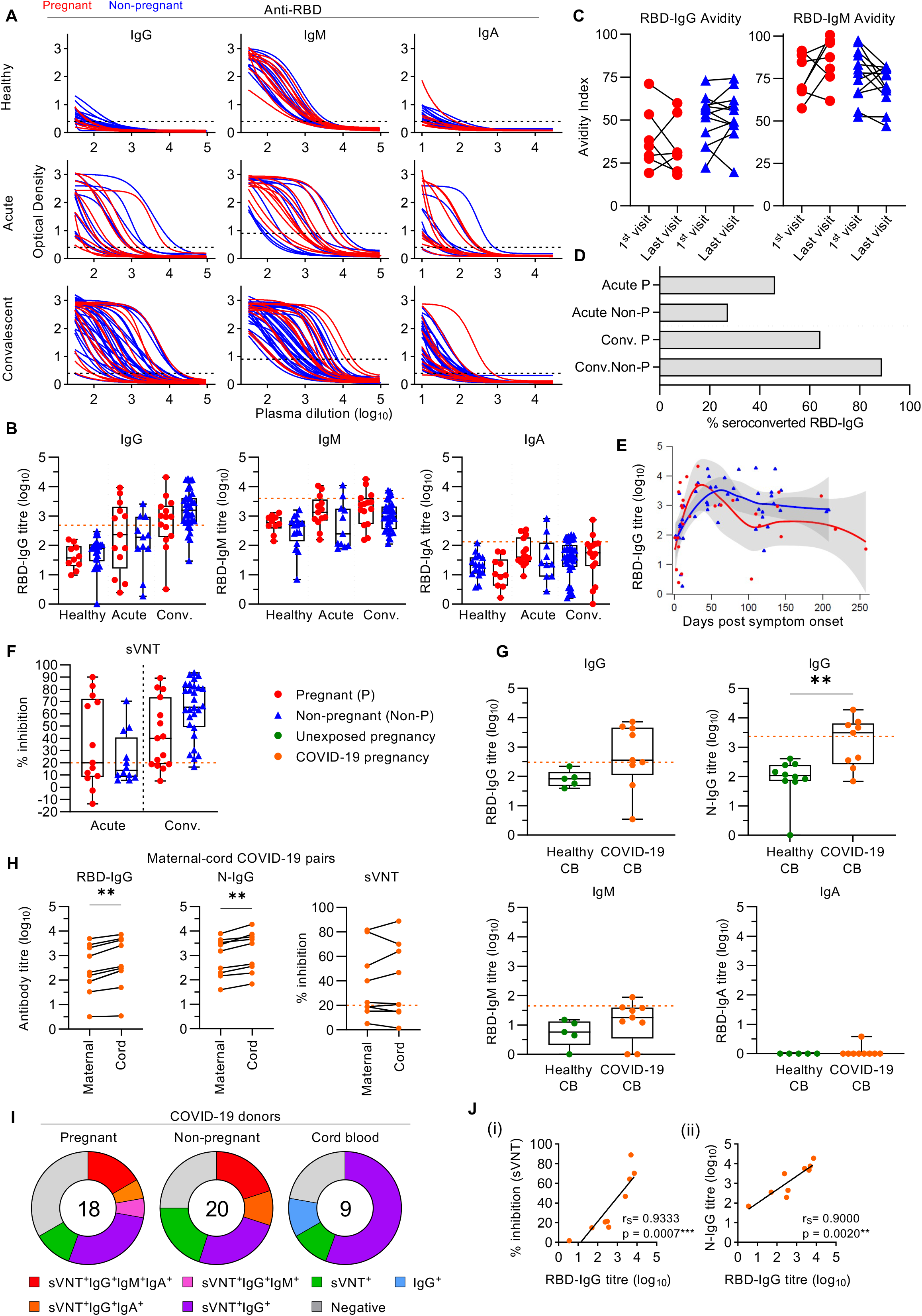
Analysis of SARS-CoV-2-specific antibodies in pregnant women and cord blood. (A) RBD-specific IgG, IgM and IgA dilution curves for healthy (P n=10, Non-P n=27X), acute COVID-19 (P n=13, Non-P n=11) or convalescent COVID-19 (P n=15, Non-P n=33X) pregnant and non-pregnant donors. (B) Log_10_ RBD-specific antibody titres in pregnant and non-pregnant donors. Orange dashed lines indicate seroconversion cut-off based on healthy pregnant and non-pregnant titres (mean plus two standard deviations). (C) Avidity of RBD-specific IgG and IgM antibodies in pregnant (n=7) and non-pregnant (n=12) COVID-19 donors performed across sequential bleeds. (D) Proportions of pregnant and non-pregnant donors who seroconverted according to RBD-specific IgG, IgM and IgA. (E) RBD-IgG kinetics for pregnant (red) and non-pregnant (blue) COVID-19 donors. LOESS regression line and 95% CI are shown. Statistics shown are Spearman correlation coefficients, ^**^*p* < 0.01. (F) Percent inhibition as determined by surrogate virus neutralization test in acute (P n=13, Non-P n=11), and convalescent (P n=15, Non-P n=28) pregnant or non-pregnant women. Orange dashed line indicates seroconversion with neutralizing antibodies. (G) Log_10_ RBD- and N-specific antibody titres in cord blood from healthy (n=5-10) and COVID-19 (n=9) pregnancies. Orange dashed lines indicate seroconversion cut-off based on healthy cord blood titres (mean plus two standard deviations). (H) RBD-specific IgG, IgM and IgA, and N-specific IgG as well as sVNT percentage inhibition in matched maternal-cord dyads (n=9). Wilcoxon test was used to determine statistical significance,^**^*p*<0.01. (I) Proportions of pregnant, non-pregnant and cord blood donors who seroconverted with different combinations of RBD-specific or neutralizing antibodies. Seroconversion was counted if a donor was positive at any timepoint if multiple samples were collected. (J) Correlations between RBD-IgG titres and (i) percent inhibition determined by sVNT or (ii) N-IgG titres in COVID-19 cord blood (n=9). Statistical significance was determined using Spearman’s rank correlation.

### Neutralizing antibodies, RBD- and N-specific IgGs cross placenta into cord blood

Cord blood plasma from COVID-19 and SARS-CoV-2 unexposed pregnancies were assessed for neutralizing antibodies as well as RBD- and nucleocapsid (N)-specific IgM, IgG and IgA antibodies. While N-specific IgG titres were significantly increased in COVID-19 cord blood compared to healthy pregnancy cord blood, with 55.5% (n=5) of donors seroconverting (Fig 2G), 66.6% (n=6) of COVID-19 cord blood donors seroconverted for RBD-IgG (Fig 2G). sVNT showed that the proportion of ACE2-RBD inhibition was similar between pregnant COVID-19 (mean 35.9% inhibition), non-pregnant COVID-19 (mean 42.8% inhibition) and COVID-19 cord blood (mean 38.2% inhibition) (Fig 2H). Similar proportions of pregnant and non-pregnant donors were positive for the combined detection of RBD-specific antibodies and ACE2-RBD inhibition (27.8% pregnant and 25.0% non-pregnant), with the majority of cord blood having RBD-IgG and/or neutralizing antibodies (77.8%) (Fig 2I).

Comparisons between matched maternal-cord dyads clearly demonstrated that RBD- and N-specific IgG were elevated in cord blood plasma (Fig 2H), which may be related to the preferential transfer of fucosylated IgG to cord blood (Atyeo et al., 2021). RBD-specific IgM and IgA were also assessed in the cord blood (Supp Fig 1A), however, as these isotypes do not vertically transfer to the fetus, it was expected that these titres were significantly lower in the cord blood compared to maternal blood. Within COVID-19 pregnancy cord blood, RBD-IgG titres strongly correlated with RBD-ACE2 inhibition (r_S_=0.9333 p=0.0007) determined by sVNT, and with N-IgG titres (r_S_=0.9000, p=0.0020) (Fig 2J). Our findings verify reports by others that SARS-CoV-2 IgG antibodies (Atyeo et al., 2021; Edlow et al., 2020; Flannery et al., 2021) and neutralizing antibodies (Joseph et al., 2021; Malshe et al., 2021) cross the placenta, providing a layer of immunity against SARS-CoV-2 infection to the neonate.

Assessment of neutralizing antibodies by microneutralization assay confirmed that there were no differences between pregnant and non-pregnant women with acute or convalescent COVID-19 (Supp Fig 1B), and that neutralizing antibodies were detected in cord blood if the matched maternal blood was also positive (Supp Fig 1C).

Differential glycosylation patterns on total IgGs, based on the number of galactose (G), sialic acid (S) and fucose (F) glycans, were detected in pregnant women, with G2S1F and G2F being significantly increased, while G0F, G1 and G1F were significantly reduced when comparing to non-pregnant women, with and without COVID-19 (Supp Fig 2). COVID-19 pregnancy cord blood had increased G2 and decreased G2S1F compared to unexposed pregnancy cord bloods. (Edlow et al., 2020)

Overall, our in-depth analysis of antibody responses in COVID-19 pregnant women clearly demonstrated generation and persistence of RBD-specific IgG, IgM and IgA antibodies in pregnant women, their SARS-CoV-2 neutralisation activity, as well as provided evidence for RBD- and N-specific IgG antibodies found in the cord blood of convalescent mothers.

### Systems serology reveals distinct antibody and Fcγ receptor profiles between pregnant and non-pregnant women

To further characterize in-depth SARS-CoV-2 specific antibody responses in pregnant and non-pregnant women as well as cord blood, a 102-parameter multiplex bead array was performed as previously described (Selva et al., 2021). A range of SARS-CoV-1 and SARS-CoV-2 spike and nucleocapsid antigens were used for the detection of a range of epitope-specific antibody subclasses and isotypes (IgG1-4, IgA1-2 and IgM) and Fcγ receptor binding (FcγRIIaH, FcγRIIaR, FcγRIIb, FcγRIIIaV, FcγRIIIaF) (Supp Table 3). To determine the key features driving the separation between two groups, data were normalized before performing a LASSO-penalized logistic regression feature selection. To classify the individuals based on the selected features, principal component analysis (PCA) was performed.

A comparison between pregnant and non-pregnant women with acute COVID-19 revealed a clear separation between groups on PC1 (27.88%) (Fig 3Ai). We identified fourteen features that contributed to the difference between pregnant and non-pregnant women with acute SARS-CoV-2 infection, twelve of which were biased towards the pregnant group with increased SARS-CoV-2 spike-head-specific IgA2, SARS-CoV-1 trimeric-spike-specific IgA1, IgA2, IgG2 and IgG4, SARS-CoV-1 nucleocapsid (NP)-specific IgA1, SARS-CoV-2 trimeric-spike-specific IgG1 and IgM, RBD-specific IgM and IgG2, spike-stalk-specific IgG3 and FcγRIIIaV (Fig 3Aii). Whereas, non-pregnant women had increased SARS-CoV-2 trimeric-spike-specific FcγRIIb and SARS-CoV-1 trimeric-spike-specific IgG3 (Fig 3Aii).

**Figure 3.**
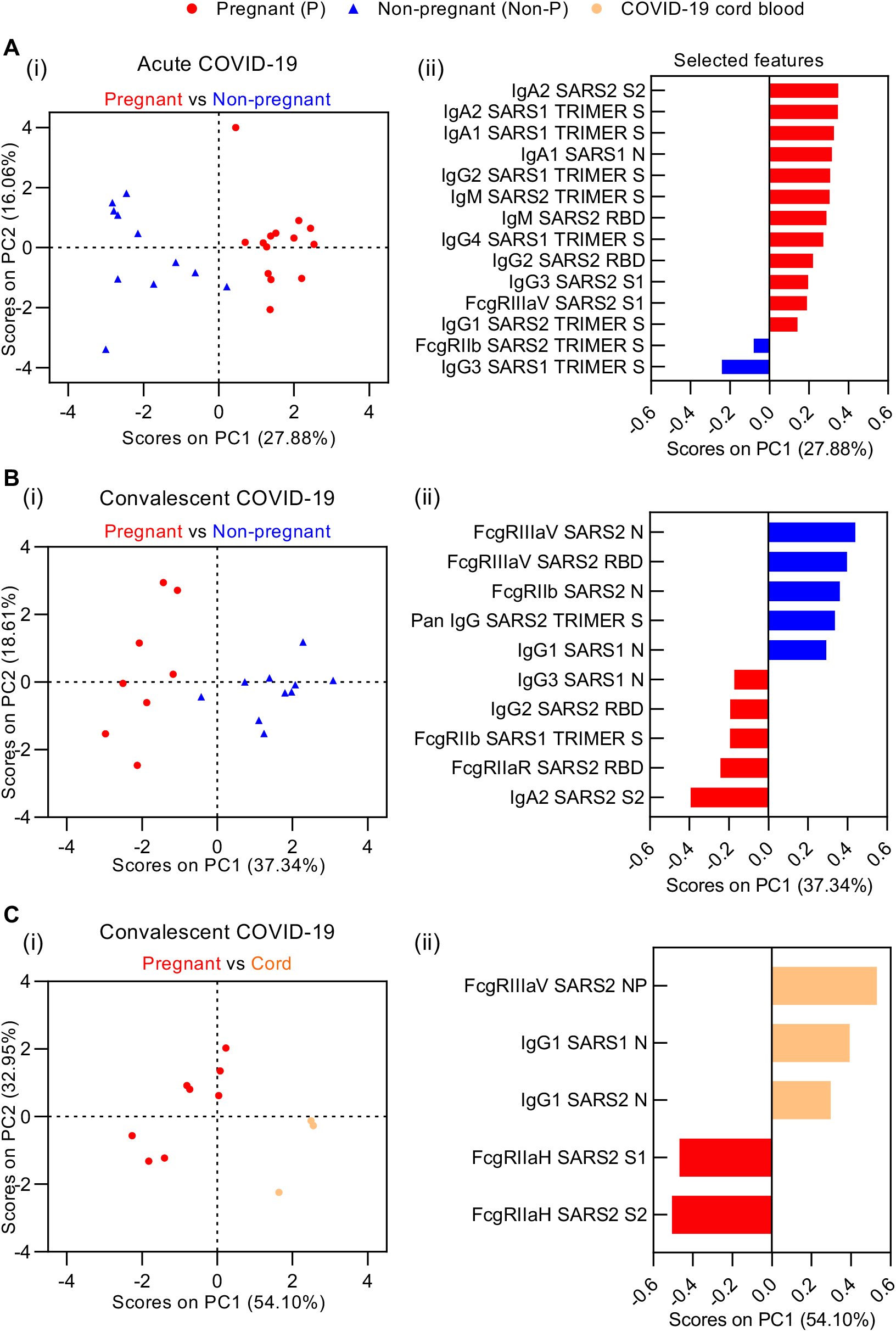
Multiplex analysis of antibody subclasses and isotypes, and FcγRs. (Ai) Principal component plots showing pregnant (red, n=13) and non-pregnant (blue, n=11) donors with acute COVID-19. (Aii) Loading plot showing the 14 selected features that cause pregnant and non-pregnant donors to separate along the PC1 axis. Principal component plot of selected features. (Bi) Principal component plots showing pregnant (red, n=8) and non-pregnant (blue, n=10) donors with convalescent COVID-19. (Bii) Loading plot showing the 10 selected features that cause pregnant and non-pregnant donors to separate along the PC1 axis. Principal component plot of selected features. (Ci) Principal component plots showing convalescent pregnant (red, n=8) and cord blood (orange, n=3) from COVID-19 pregnancies. (Cii) Loading plot showing the five selected features that cause pregnant and non-pregnant donors to separate along the PC1 axis. Principal component plot of selected features. IgM and IgA features were excluded from this comparison due to the lack of these isotypes in the cord blood which would masked any antigen-specific findings. SARS2, SARS-CoV-2; SARS1, SARS-CoV-1; S, spike, S1, spike subunit 1 (stalk); S2, spike subunit 2 (head); N, nucleocapsid; RBD, receptor binding domain.

To understand whether the serology features changed over time, we subsequently performed PCA on convalescent pregnant and non-pregnant donors (Fig 3B). Ten features were found to drive the difference between pregnant and non-pregnant women at convalescence (Fig 3Bii) and provided 37.34% variance of groups on PC1 (Fig 3Bi). Among the features elevated in convalescent non-pregnant women, SARS-CoV-2 nucleocapsid-specific FcγRIIIaV and FcγRIIb, RBD-specific FcγRIIIaV, spike-trimer-specific total IgG and SARS-CoV-1 nucleocapsid-specific IgG1 dominated (Fig 3Bii). In contrast, pregnant women displayed more spike-stalk-specific IgA2, SARS-CoV-2 RBD-specific IgG2 and FcγRIIaR, spike-trimer-specific FcγRIIb and SARS-CoV-1 nucleocapsid-specific IgG3.

As IgG, but not IgM or IgA, is transferred across the placenta during gestation, our feature selection model of convalescent pregnant women and cord blood from COVID-19 pregnancies excluded IgM and IgA parameters to accurately reveal differences in virus-specific antibodies rather than their subclasses. PCA demonstrated that convalescent pregnant women and COVID-19 pregnancy cord blood were separated across PC1 (Fig 3Ci, Variance 54.10%). Cord blood from COVID-19 pregnancies were enriched for IgG1 and FcγRIIIaV against SARS-CoV-1 or SARS-CoV-2 nucleocapsid protein, while convalescent pregnant women had elevated SARS-CoV-2 spike stalk and head-specific antibodies with the capacity to bind FcγRIIaH (Fig 3Cii). Thus, our system serology approach revealed distinct antibody and Fcγ receptor profiles between pregnant and non-pregnant women, as well as between pregnant women and COVID-19 cord blood.

### Similar frequencies of antibody-secreting cells, circulating T_FH_ cells and monocytes in pregnant and non-pregnant COVID-19

As B cells are needed for antibody production, we determined B cell activation phenotypes by flow cytometry. Analysis of antibody-secreting cells (ASCs) defined as CD27^+^CD38^+^ of the CD19^+^CD3^-^ lymphocyte population (Fig 4A) showed comparable frequencies between pregnant and non-pregnant women with acute (mean 5.5% and 3.7%, respectively) and convalescent (mean 1.0% and 0.8%, respectively) COVID-19 (Fig 4B). However, the mean fold-difference in the frequency of ASCs at the acute phase compared to healthy individuals was ∼6-fold higher in pregnant women compared to a lower ∼2-fold difference in non-pregnant women (Fig 4C), although ASCs displayed similar kinetics in both patient groups (Fig 4D). RBD-specific antibody titres and the frequency of ASCs did not correlate in either pregnant or non-pregnant women with acute or convalescent COVID-19 (Fig 4E), similar to previous findings (Koutsakos et al., 2021; Kuri-Cervantes et al., 2020; Mathew et al., 2020).

**Figure 4.**
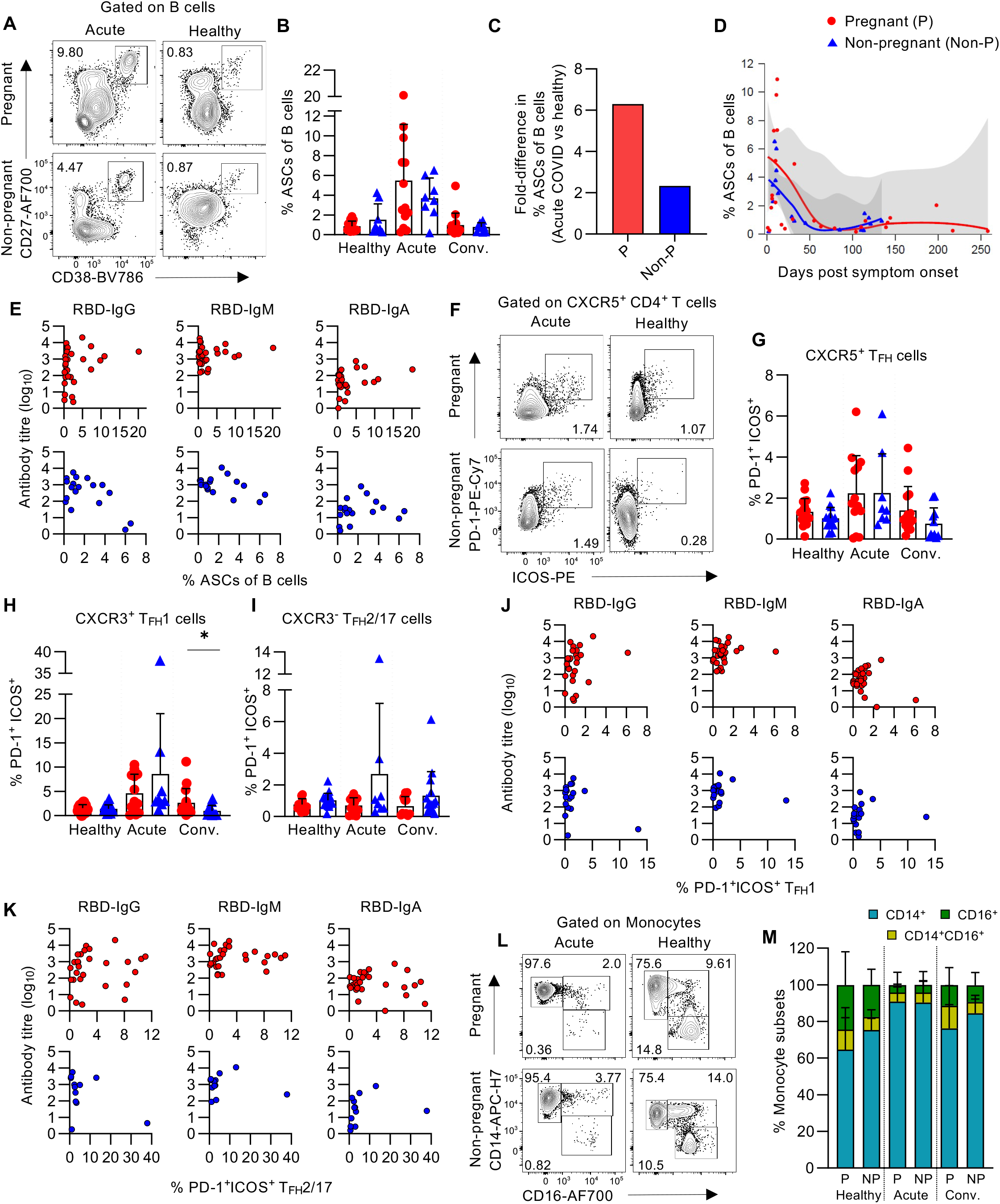
Activation in antibody-secreting B cells and CD4^+^ T follicular helper cells. (A) CD27^+^CD38^+^ antibody-secreting cells (ASCs) were gated from the CD19^+^CD3^-^ B cell population. (B) Frequencies of ASCs of B cells in healthy (P n=18, Non-P n=8), acute (P n=13, Non-P n=8) or convalescent (P n=15, Non-P n=10) pregnant or non-pregnant women. Means and standard deviations are shown. (C) Fold-difference in the mean frequency of ASCs between healthy and acute COVID-19 for pregnant and non-pregnant donors. (D) Kinetics of ASCs against the day post symptom onset for pregnant and non-pregnant COVID-19 donors. LOESS regression line and 95% CI are shown. (E) Correlations between RBD-specific antibody titres and the frequency of ASCs for pregnant and non-pregnant COVID-19 donors. No statistical significance was reached when using the Spearman correlation method. (F) T_FH_ cells were defined as CXCR5^+^CD4^+^ T cells and their activation was determined by expression of PD-1 and ICOS. (G-I) Frequencies of PD-1^+^ICOS^+^ total T_FH_ cells (G), CXCR3^+^ T_FH_1 cells (H) and CXCR3^-^ T_FH_2/17 cells (I) healthy (P n=18, Non-P n=14), acute (P n=13, Non-P n=8) or convalescent (P n=15, Non-P n=11) pregnant or non-pregnant women. (J-K) Correlations between RBD-specific antibodies and PD-1^+^ICOS^+^ T_FH_1 or T_FH_2/17 (K) cells. No statistical significance was reached when using the Spearman correlation method. (L) Differential gating of CD14^+^CD16^-^ classical, CD14^+^CD16^+^ inflammatory and CD14^-^CD16^+^ patrolling monocytes. (M) Proportions of classical, inflammatory and patrolling monocytes. Means and standard deviations are shown. No statistical significance was achieved when comparing individual monocyte populations in pregnant and non-pregnant donors by Mann-Whitney *U* test.

Circulating CXCR5^+^ T follicular helper (cT_FH_) CD4^+^ T cells correlate with B cell maturation and activation (Koutsakos et al., 2018), therefore cT_FH_ cells were assessed by flow cytometry, with activation defined as co-expression of PD-1 and ICOS (Fig 4F). No significant differences in the frequencies of PD-1^+^ICOS^+^ cT_FH_ cells were observed between pregnant and non-pregnant women with acute or convalescent COVID-19 (Fig 4G). When cT_FH_ cells were further defined into CXCR3^+^ T_FH_ type 1 (T_FH_1) and CXCR3^-^ T_FH_ type 2/17 (cT_FH_2/17), the frequency of PD-1^+^ICOS^+^-activated cT_FH_1 cells was higher in convalescent pregnant versus non-pregnant women (mean 2.63% vs 0.96%, respectively, *p*<0.05), however no differences were found for the acute or healthy groups (Fig 4H). Similar activation levels of the cT_FH_2/17 subset were observed in pregnant and non-pregnant women for each of the disease states (Fig 4I). There were no correlations between RBD-specific antibody titres and the frequency of activated cT_FH_1 or cT_FH_2/17 cells (Fig 4J,K).

Analysis of the myeloid compartment revealed no differences in monocyte subsets according to their CD14 and/or CD16 expression across healthy, COVID-19 acute or convalescent pregnant and non-pregnant women (Fig 4L,M). Therefore, our results highlight that pregnant woman can generate robust ASC, T_FH_ and myeloid responses during acute COVID-19.

### Differential NK cell activation and cytotoxicity patterns in pregnant women during acute COVID-19

Natural killer cells play an important role in anti-viral immunity, especially via mediating rapid killing of virally-infected cells. To determine activation of NK cells, CD3^-^CD56^+^ NK cells were assessed for upregulation of HLA-DR^+^ expression (Fig 5A). In healthy state, total NK cell activation was significantly higher in healthy pregnant women when compared to non-pregnant women (mean 4.49% and 0.2%, respectively, *p*<0.0001) consistent with previous work (Le Gars et al., 2019), indicating pre-activated NK cells during healthy pregnancy. Strikingly, activation profiles of these pre-activated NK cells remained unchanged at acute and convalescent COVID-19 in pregnancy (Fig 5B). In contrast to non-pregnant COVID-19 women, where NK cell activation during acute COVID-19 was driven by a ∼28-fold increase in NK cell activation (as compared to healthy non-pregnant), NK activation in acute COVID-19 pregnant women remained at the level observed in healthy pregnant participants (Fig 5C). Conversely, at convalescence, while the proportion of activated NK cells decreased in non-pregnant women with COVID-19, NK cell activation levels still remained high for at least >100 days post disease onset in convalescent COVID-19 pregnant women, similar to the high levels already observed for healthy and acute pregnant women. This was further evident from a significant negative correlation between the frequency of HLA-DR^+^ NK cells and the day post symptom onset (r_s_=-0.5496, *p*=0.0294) in non-pregnant women, with no correlation observed in pregnant COVID-19 women (Fig 5D). Overall, our data reveal pre-activated state of NK cells in healthy pregnancy as well as tightly regulated processes of NK activation above this pre-activated level, resulting in lack of further NK cell activation during acute COVID-19 in pregnancy.

**Figure 5.**
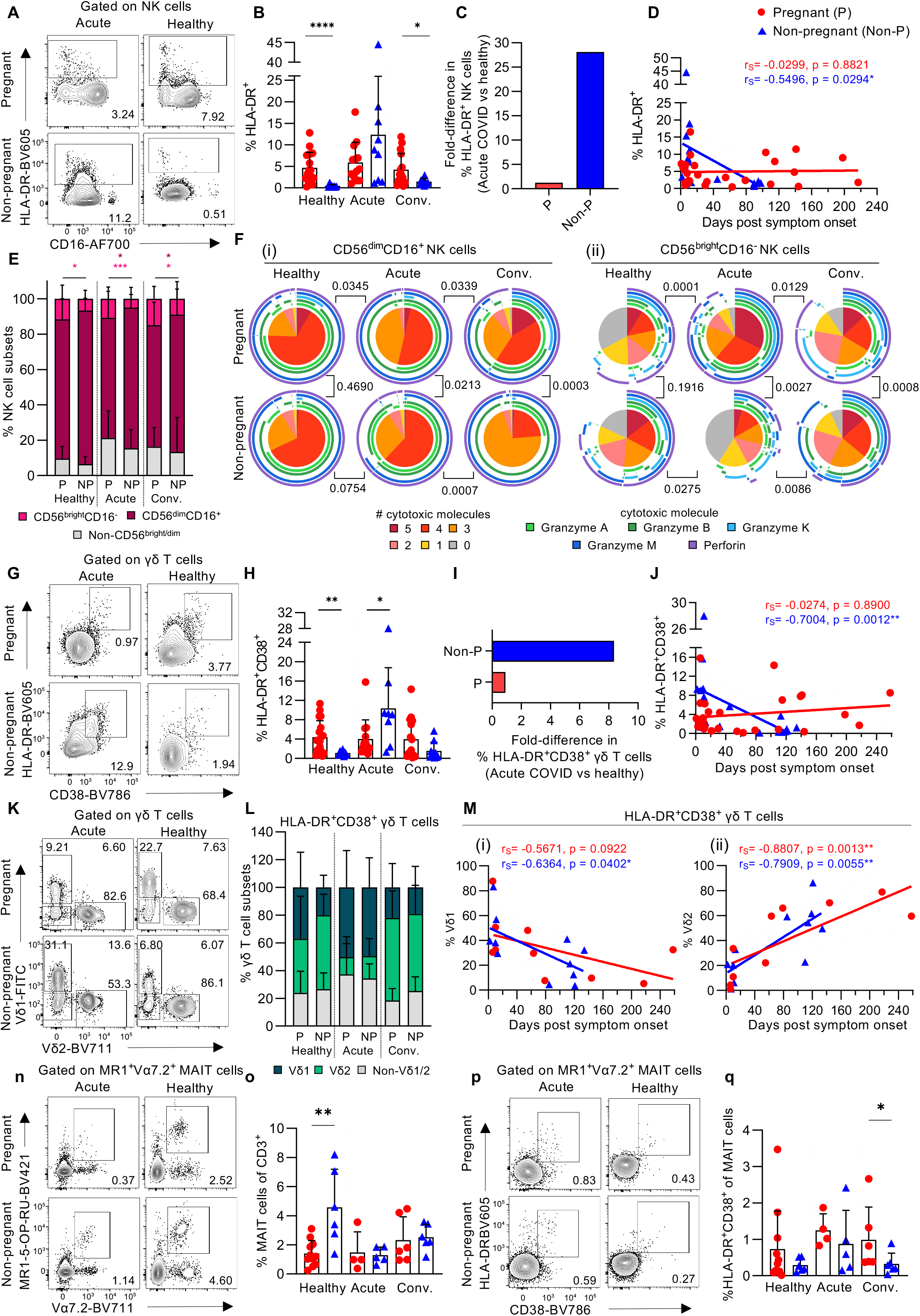
Differential NK cell and γδ T cell activation in pregnant women with COVID- 19. (A) NK cells were defined as CD3^-^CD56^+^ and their activation was determined by HLA-DR expression. (B) Frequencies of HLA-DR^+^ NK cells in healthy (P n=18, Non-P n=8), acute (P n=13, Non-P n=9) and convalescent (P n=15, Non-P n=10) COVID-19 pregnant and non-pregnant women. ^*^*p*<0.05, ^****^*p*<0.0001. (C) Fold-difference in the mean frequency of HLA-DR^+^ NK cells from healthy to acute COVID-19 for pregnant and non-pregnant donors. (D) Correlation between the frequency of HLA-DR^+^ NK cells and days post symptom onset for pregnant and non-pregnant women with COVID-19. LOESS regression line and 95% CI are shown. Statistics shown are Spearman correlation coefficient, ^*^*p*<0.05. (E) Proportions of CD56^bright^CD16^-^, CD56^dim^CD16^+^, and intermediate non-CD56^bright/dim^ NK cells. ^*^p<0.05,^***^p<0.001. (F) Proportions of granzymes A, B, K and M and perforin expression in total, (i) CD56^dim^CD16^+^ or (ii) CD56^bright^CD16^-^ NK cells. Inner pie chart describes the proportions of NK cells expressing multiple cytotoxic molecules and the outer circles depict which molecules contribute to the multifunctionality. Permutations test was used to determine statistical significance. (G) γδ T cells were defined as CD3^+^γδTCR^+^ lymphocytes and their activation determined by HLA-DR and CD38 co-expression. (H) Frequencies of HLA-DR^+^CD38^+^ γδ T cells in pregnant and non-pregnant women who were healthy (P n=18, Non-P n=8) or had acute (P n=13, Non-P n=8) or convalescent (P n=15, Non-P n=10) COVID-19.^*^*p*<0.05, ^**^*p*<0.01. (I) Fold-difference in the mean frequency of HLA-DR^+^CD38^+^ γδ T cells from healthy to acute COVID-19 for pregnant and non-pregnant donors. (J) Correlation between the frequency of HLA-DR^+^CD38^+^ γδ T cells and days post symptom onset for pregnant and non-pregnant women with COVID-19. LOESS regression line and 95% CI are shown. Statistics shown are Spearman correlation coefficient, ^**^*p*<0.01. (K) Vδ1, Vδ2 and non-Vδ1/2 subsetting of γδ T cells. (L) Proportions of Vδ1, Vδ2 and non-Vδ1/2 within the HLA-DR^+^CD38^+^ γδ T cell population. (M) Correlation between the frequency of (i) Vδ1 or (ii) Vδ2 T cells and days post symptom onset for pregnant and non-pregnant women with COVID-19. (N) MAIT cells defined by binding of MR1-5-OP-RU tetramer and expression of Vα7.2. (O) MAIT cell frequencies of CD3^+^ lymphocytes. (P) HLA-DR and CD38 expression on MAIT cells. (Q) Frequencies of HLA-DR^+^CD38^+^ MAIT cells in pregnant and non-pregnant women who were healthy (P n=11, Non-P n=6) or had acute (P n=4, Non-P n=5) or convalescent (P n=6, Non-P n=6) COVID-19. ^**^*p*<0.01. Means and standard deviations are shown. Mann Whitney *U* test was used to determine statistical significance unless otherwise stated.

To further delve into differential NK cell activation between pregnant and non-pregnant women, we differentiated the NK cell population into CD56^bright^CD16^low/-^ and CD56^dim^CD16^+^ subsets. CD56^bright^ NK cells are functionally associated with cytokine production, whereas CD56^dim^ NK cells are cytotoxic (Caligiuri, 2008; Cooper et al., 2001). In each of the healthy or COVID-19 disease states, pregnant women had a significantly higher frequency of CD56^bright^ NK cells compared to non-pregnant women, while they had significantly lower frequencies of CD56^dim^ NK cells during acute and convalescent COVID-19 (Fig 5E).

To assess the cytotoxic potential between the two CD56^bright^CD16^low/-^ and CD56^dim^CD16^+^ NK cell subsets, we performed intracellular staining for granzyme A, B, K and M as well as perforin. The majority of the total NK cell population expressed 3-5 cytotoxic granzymes or perforin in pregnant and non-pregnant women who were healthy or had acute or convalescent COVID-19. CD56^dim^ NK cells largely expressed multiple cytotoxic molecules, fitting their previously defined cytotoxic function (Fig 5Fi). Conversely, CD56^bright^ NK cells displayed less multifunctionality overall, but interestingly, pregnant women with acute COVID-19 had the largest proportions of cells expressing 3-5 cytotoxic molecules, which might indicate increased cytotoxic potential of this classically non-cytotoxic NK cell subset during pregnancy (Fig 5Fii). This might provide a partial explanation on the need for a tight regulation of highly cytotoxic NK cells during COVID-19 in pregnant women.

### Differential γδ T cell activation in pregnant women during COVID-19

γδ T cells are an unconventional T cell subset which play an important role in anti-viral responses to respiratory diseases, including influenza (Sant et al., 2019) and COVID-19 (Jouan et al., 2020), however their role and activation status in COVID-19 pregnant women is not yet defined. We defined activation of γδ T cells by HLA-DR and CD38 co-expression on CD3^+^γδTCR^+^ lymphocytes from peripheral blood (Fig 5G). Similar to our findings in NK cells, the proportion of activated HLA-DR^+^CD38^+^ γδ T cells was higher in healthy pregnant women compared to non-pregnant women (mean 4.4% and 1.1%, respectively, *p*<0.01; Fig 5H). Alternatively, during acute COVID-19, pregnant women had lower frequencies of activated γδ T cells than non-pregnant women (mean 4.0% and 10.4%, respectively, *p*<0.05; Fig 5H). The activation of γδ T cells in pregnant women remained stable despite having acute COVID-19, whereas non-pregnant women with acute disease displayed an 8-fold increase in γδ T cell activation compared to healthy non-pregnant women (Fig 5I). The frequency of HLA-DR^+^CD38^+^ γδ T cells had a significant negative correlation with the day post symptom onset in non-pregnant women (r_s_=-0.7004, *p*=0.0012), however these two variables did not correlate for pregnant women (Fig 5J). Further probing of HLA-DR^+^CD38^+^ γδ T cells revealed a similar distribution in the activation of the Vδ1 or Vδ2 subsets between pregnant and non-pregnant women (Fig 5K,L). However, during acute COVID-19, we observed that the Vδ1 subset comprised a larger proportion of activated γδ T cells, which then decreased at convalescence for pregnant and non-pregnant women (Fig 5M). As Vδ2 T cells are associated with their cytotoxic activity (Juno and Kent, 2020; Wragg et al., 2020), the decreased proportion in the blood during acute COVID-19 could be due to trafficking to the site of infection to perform effector functions.

It is important to note that while key differences in the activation of non-classical γδ T cells and NK cells were observed, similar activation patterns of classical αβ CD4^+^ and CD8^+^ T cell activation were detected in COVID-19 pregnant and non-pregnant women, suggesting that T cell receptor (TCR)-mediated T cell responses in the context of peptide/MHC presentation might not be affected by pregnancy (Supp Fig 3).

### Regulated activation of mucosal-associated invariant T (MAIT) cell responses in pregnant women during COVID-19

Defined by their ability to recognize MR1 and expression of the Vα7.2 TCR chain, MAIT cells represent another subset of unconventional T cells and have an important role in anti-viral immunity, such as during influenza virus infection and COVID-19 (Flament et al., 2021; Jouan et al., 2020; Loh et al., 2016). While MAIT cells characteristically recognize riboflavin metabolites produced by bacterial biosynthesis pathways, they can be activated as a first-line of defence in a TCR-independent manner through cytokines, including IL-12 and IL-18 stimulation (Loh et al., 2016; Mayassi et al., 2021). Our analysis revealed that healthy pregnant women had a lower frequency of MR1-5-OP-RU tetramer^+^ Vα7.2^+^ MAIT cells compared to healthy non-pregnant women (Fig 5N,O). Non-pregnant women displayed a decrease in the frequency of MAIT cells during acute COVID-19, however a further reduction from the healthy state was not observed in pregnant women, likely due to the already perturbed MAIT cell frequencies pre-COVID-19 (Fig 5O). However, further investigation of MAIT cell activation defined by the expression of HLA-DR and CD38 showed unchanged activation phenotype in pregnant women across disease stages, while non-pregnant women displayed increased activation during acute COVID-19 (Fig 5P,Q).

### Immune cell activation within the placenta is similar between COVID-19 convalescent and unexposed pregnancies

The placenta is the organ that forms the maternal-fetal interface and allows for exchange of nutrients and waste products to and from the fetus. There is limited evidence that SARS-CoV-2 is able to cross the placenta, resulting in vertical transmission of the virus(Fenizia et al., 2020; Kotlyar et al., 2021). To assess for any lasting changes in cellular immune components within placental tissue, flow cytometry was performed on placenta single-cell suspensions from 9 COVID-19 convalescent and 6 SARS-CoV-2 unexposed pregnancies. There was no difference in total NK cell activation between COVID-19 and unexposed placenta samples (Supp Fig 4A), and a similar finding was observed in each of the CD56^bright^ and CD56^dim^ subsets (Supp Fig 4B,C). Contrary to observations in maternal blood at both acute and convalescent timepoints (Fig 5E), there were almost equivalent proportions of CD56^bright^ and CD56^dim^ subsets comprising the placental NK cell population for healthy (mean 34.8% and 45.1%, respectively) and COVID-19 convalescent pregnancies (mean 41.2% and 33.9%, respectively, Supp Fig 4D). There were significantly higher frequencies of CD56^bright^ NK cells in the placenta tissue compared to matched maternal blood (mean 13.4% and 38.8%, *p*=0.0005, Supp Fig 4E), typically observed in healthy pregnancies (Liu et al., 2021b). αβ CD4^+^ and CD8^+^ T cells and γδ T cells were also examined in the placenta for HLA-DR and CD38 expression to determine their activation. Similar to NK cells, there were no significant differences in the activation of these T cell subsets (Supp Fig 4F-H), indicating no lasting differential activation within the placenta at convalescence from COVID-19.

### Elevated levels of IL-1β, IFN-γ, IL-8, IL-18 and IL-33 in healthy pregnancy and during COVID-19

As dysregulation of inflammatory cytokines and chemokines can be associated with severe COVID-19 (Koutsakos et al., 2021; Zhang et al., 2020), we assessed cytokine and chemokine profiles in our pregnancy cohort to understand whether the inflammatory response during COVID-19 differed between pregnant and non-pregnant women. As pregnant women are in a differential state of inflammation due to gestation, with relatively higher levels of IL-1β, IL-4, IL-5 and IL-10 (Pinheiro et al., 2013), we hypothesised that this might impact cytokine and chemokine levels during acute COVID-19. Elevated levels of IL-1β, IFN-γ, IL-8, IL-18 and IL-33 were found in pregnant women in their healthy state, and these cytokine levels remained elevated during acute and convalescent COVID-19 (Fig 6A). In addition to differences in cytokine and chemokine levels individually, the total level of cytokines and chemokines measured was higher in pregnant women during each disease state (Fig 6B). Despite having a greater total concentration of cytokines and chemokines, pregnant women with acute COVID-19 had similar proportions of each cytokine in the plasma compared to healthy and convalescent samples (Fig 6C). Moreover, similar levels of cytokines were detected in cord blood from healthy and COVID-19 convalescent pregnancies (Supp Fig 5). Therefore, our data suggest that pregnant women do not make the same inflammatory COVID-19 response as non-pregnant women, as some cytokine levels are already high in healthy pregnancy, they fail to rise further during acute COVID-19 and then fail to decrease following SARS-CoV-2 infection.

**Figure 6.**
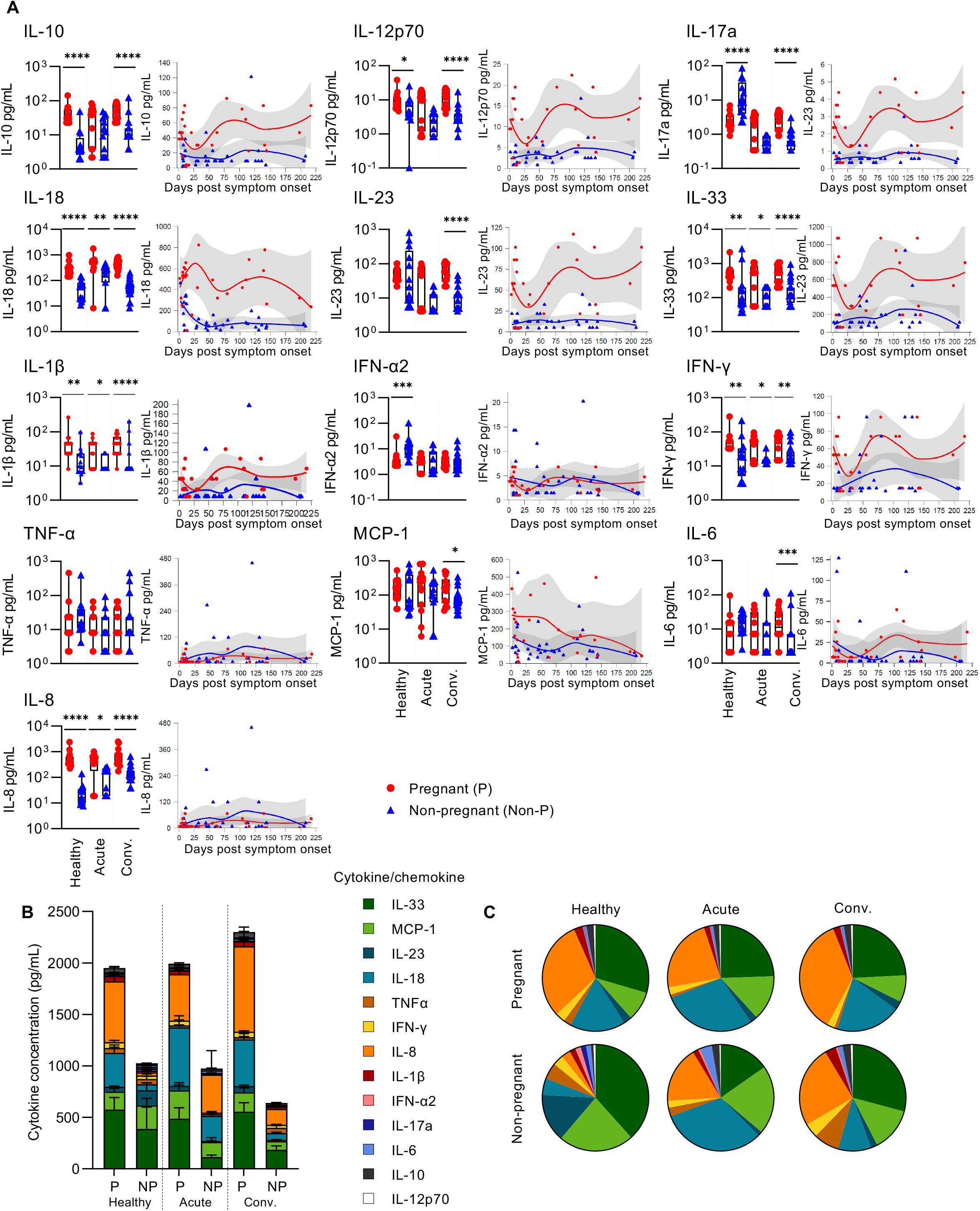
Cytokine and chemokine concentrations and proportions within blood plasma. (A, left) Concentrations of 13 cytokines and chemokines measured in pregnant and non-pregnant women who were healthy (P n=15, Non-P n=11) or had acute (P n=13, Non-P n=11) or convalescent (P n=14, Non-P n=26) COVID-19. Mann Whitney *U* test was used to determine statistical significance, ^*^*p*<0.05, ^**^*p*<0.01, ^***^*p*<0.001, ^****^*p*<0.0001. (A, right) Correlation between cytokine or chemokine concentration and days post symptom onset for pregnant and non-pregnant COVID-19 donors. LOESS regression line and 95% CI are shown. Statistics shown are Spearman correlation coefficient, ^*^*p*<0.05. (B) Quantification of the total cytokine and chemokine concentration. Means and standard deviations are shown. (C) Mean proportion of each cytokine and chemokine in pregnant and non-pregnant women who were healthy or had acute or convalescent COVID-19.

### Immune network analysis reveals a comprehensive map of immune responses to COVID-19 in pregnancy

We comprehensively analysed all the immunological parameters between pregnant and non-pregnant women, which included 217 datasets of antibodies, cellular subsets and cytokines/chemokines and revealed distinct profiles between pregnant and non-pregnant women (Fig 7A). While striking differences in immune responses were found between pregnant and non-pregnant women in the healthy state and convalescent COVID-19, immune responses were more comparable during acute COVID-19 (Fig 7B-D), which reflects the pre-activated and inflamed state in pregnant women. For example, in the healthy state, pregnant women displayed profound upregulation of HLA-DR^+^CD56^bright^ NK cells, HLA-DR^+^CD38^+^ γδ T cells, IL-8, IL-10, and IL-18 (Fig 7B). These differences became less apparent during acute COVID-19, when pregnant women appeared to have prototypical antiviral immunity (Fig 7C). Thus, although the immune responses in pregnant women with acute COVID-19 closely resemble those in acute COVID-19 non-pregnant women, a lack of activation above the healthy baseline levels questions the quality of these responses. Conversely, at convalescence, hyperactivation of immune responses in pregnant women was again observed, especially with respect to cytokine production (IL-8, IL-10, IL-12, IL-17a, IL-18, IL-23) and NK cell cytotoxicity (Fig 7D). Taken together, our study provides a comprehensive map of longitudinal immunological responses in COVID-19 pregnant women during acute and convalescent phases of SARS-CoV-2 infection and reveals pre-activated status of NK cells and unconventional T cells and elevated levels of cytokines in healthy pregnancy, which remained unchanged during acute and convalescent COVID-19.

**Figure 7.**
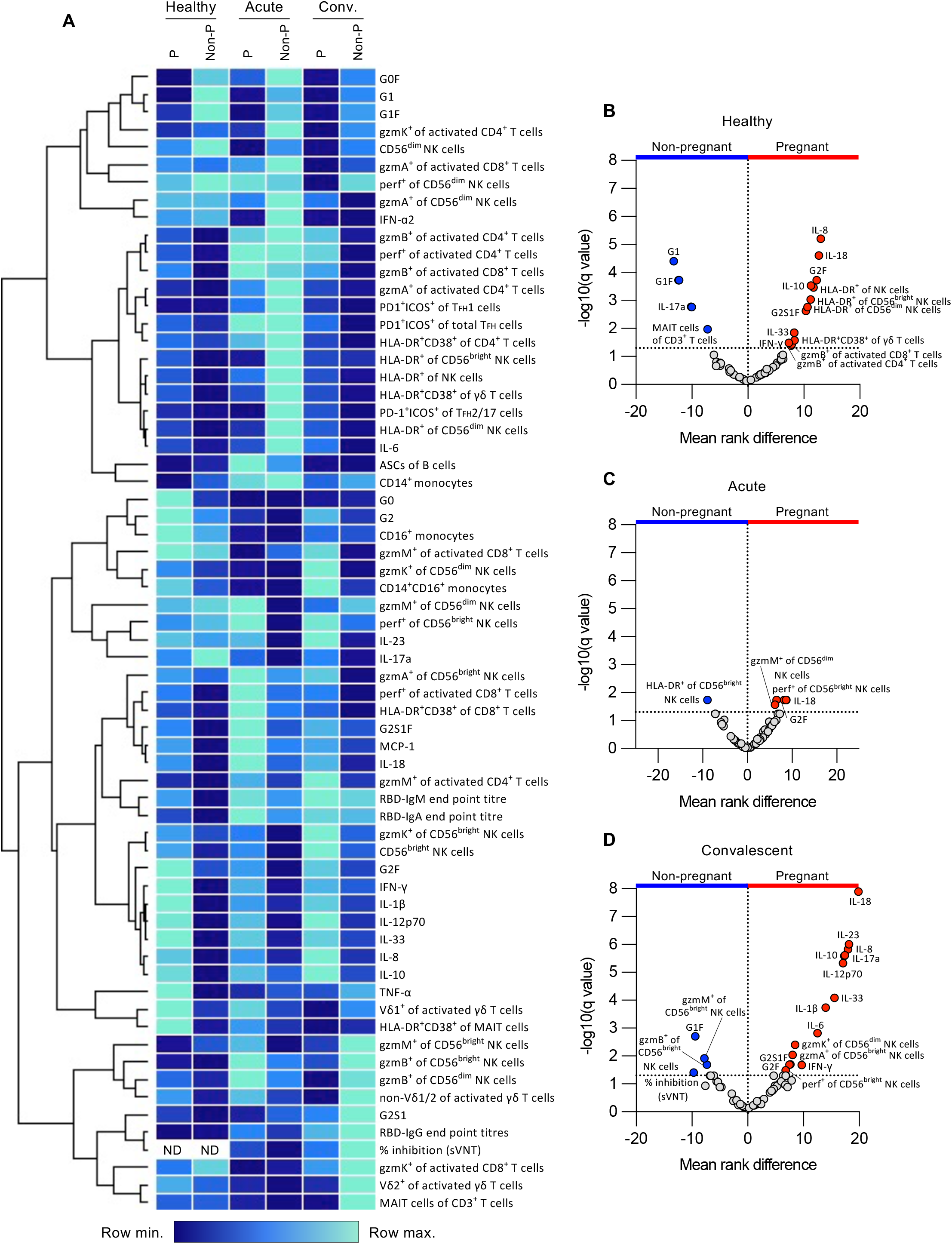
Summary data of key differences in immune parameters between pregnant and non-pregnant women. (A) Heatmap depicting the mean of 65 selected immune parameters for healthy, acute and convalescent pregnant and non-pregnant women. Data shown are calculated from z-scored values. (a-c) Volcano plots of 65 selected cellular and humoral immune parameters between pregnant and non-pregnant women who were (B) healthy or had (C) acute or (D) convalescent COVID-19.

## DISCUSSION

Pregnant women are considered a vulnerable group for SARS-CoV-2 infection (Zambrano et al., 2020), as published reports correlated pregnancy with an increased risk of ICU admission, invasive ventilation, extracorporeal membrane oxygenation (ECMO) (Allotey et al., 2020; Zambrano et al., 2020), death, sepsis, mechanical ventilation, ICU admission, shock, acute renal failure and thromboembolic disease (Ko et al., 2021) and hypertensive complications (Papageorghiou et al., 2021). However, not all studies reveal strong correlations between pregnancy and COVID-19 severity or prolonged disease complications (Crovetto et al., 2020), and not at the level that occurred during the 2009 H1N1 pandemic (Siston et al., 2010). As more epidemiological data regarding COVID-19 during pregnancy emerges, it is essential to comprehensively define immune responses to SARS-CoV-2 infection to understand whether SARS-CoV-2 immunity in pregnancy is prototypical and resembles immune responses of mild to moderate COVID-19 in non-pregnant individuals (Juno et al., 2020c; Koutsakos et al., 2021; Rowntree et al., 2021; Thevarajan et al., 2020; Wheatley et al., 2021), or in contrast, is characterised by immune perturbations similar to those observed during severe COVID-19 (Koutsakos et al., 2021; Lucas et al., 2020). In-depth dissection of immune responses in pregnant women is needed to provide insights into the immunological basis underlying COVID-19 outcomes. Our study fills this knowledge gap and provides the first comprehensive map of longitudinal immunological responses in COVID-19 pregnant women during acute and convalescent phases of SARS-CoV-2 infection. We investigated the breadth of 217 cellular and humoral immune parameters in pregnant women, as well as immune responses in cord blood and placenta from COVID-19 pregnancies. To the best of our knowledge, this is the first study providing an integrated comprehensive dataset on innate, adaptive and cellular immune networks to SARS-CoV-2 infection in pregnant women.

Importantly, our in-depth analysis of the antibody response in COVID-19 pregnant women via elucidating RBD-specific IgG, IgM and IgA, their neutralising activity, multidimensional system serology parameters together with ASCs and T_FH_ cells showed comparable antibody features between COVID-19 pregnant and non-pregnant women during acute and convalescent SARS-CoV-2 infection. Our analyses of humoral immune responses to SARS-CoV-2 in pregnancy clearly demonstrated generation and persistence of RBD-specific IgG, IgM and IgA antibodies in pregnant women, and their SARS-CoV-2 neutralisation activity, together with rapid induction of ASC and T_FH_ cells. We also provided evidence of RBD- and N-specific IgG antibodies found in the cord blood of convalescent mothers. Our data validates previous studies showing generation of SARS-CoV-2-specific antibodies in pregnant women (Atyeo et al., 2021; Edlow et al., 2020; Jang et al., 2021; Wang et al., 2021).

Our longitudinal comparisons of cellular immunity in COVID-19 revealed prototypical activation patterns of classical αβ CD4^+^ and CD8^+^ T cells but differential and NK cell and unconventional T cell activation in pregnant women during acute COVID-19, with NK cells expressing increased frequency of CD56^bright^ NK cells and multiple cytotoxic molecules. Differential immune responses by NK cells have been previously reported in the literature in healthy pregnant women and in the context of influenza(Kay et al., 2014; Le Gars et al., 2019), however γδ T cell and MAIT activation in pregnancy is not well understood. It was recently reported that healthy pregnant women in their second or third trimester have increased frequencies of a γδ T cell subset characterised by CD56 expression and higher cytotoxic potential reflected by expression of CD107a, in comparison to non-pregnant women(Nörenberg et al., 2021). Additionally, women who were previously pregnant had substantially increased frequencies of PD-1^+^ Vδ2^+^ γδ T cells compared to nulliparous women or women with recurrent pregnancy loss (Liu et al., 2021a). Furthermore, an enrichment of Vδ1 and HLA-DR^+^ γδ T cells have been observed in the decidua during early pregnancy(Terzieva et al., 2019). These reports suggest an important role for γδ T cells in pregnancy, but their function and mechanisms in this state remain unknown.

While striking differences in MAIT and γδ T cell, and NK cell responses were found between pregnant and non-pregnant women in healthy and/or convalescent COVID-19 states, immune responses appeared reasonably comparable during acute COVID-19. As a result, there was no increased activation of γδ T cell and NK cell above the ‘baseline’ healthy pregnant levels, and no further decrease in the frequencies of peripheral blood MAIT cells. As anti-viral immune responses are multifactorial, further studies performed in γδ T cell and NK cell knockout mice during SARS-CoV-2 infection are needed to fully understand the impact of differential NK cell and γδ T cell activation in the context of COVID-19.

IL-1β, IFN-γ, IL-8, IL-18 and IL-33 were increased in healthy pregnant women and during acute and convalescent COVID-19. Interestingly, IL-18 dependent MAIT cell activation has been reported in the context of influenza infection (Loh et al., 2016), which might suggest a role for IL-18 in mediating MAIT cell activation during pregnancy as it was found at increased concentrations in our analysis. There are limited studies that compare cytokine levels between healthy pregnant and non-pregnant women to define a common cytokine profile associated with pregnancy. However, Pinheiro et al. (2013) showed that pregnant women have an increased frequency of regulatory cytokines, including IL-1β, IL-4, IL-5 and IL-10, compared to non-pregnant women. Furthermore, in a comparison of cytokine levels during pregnancy and at one-year post-partum, Graham et al. (2017) found that pro-inflammatory IL-18, TNFα and MCP-1 were reduced during pregnancy while IL-10 remained unchanged. Pregnant women with COVID-19 shared similar cytokine and chemokine profiles as non-pregnant women, with the exception of eotaxin and GRO-a, while key differences between COVID-19 and healthy pregnant women have been observed in IL-12p70, MIP-1β, RANTES (Chen et al., 2021; Zhao et al., 2021). Our analysis showed that IL-18 was elevated during pregnancy regardless of disease state, which has been shown to cause TCR-independent activation of MAIT cells (Loh et al., 2016). It would be of value to further investigate if the pre-activated phenotype observed in MAIT cells of pregnant women could be related to the increased concentration of this cytokine in the blood plasma.

Our analyses of immune cell populations within placenta tissues from COVID-19 or healthy pregnancies revealed comparable levels of activation in NK and T cell subsets. The similarities between healthy and COVID-19 placenta immune cell activation might be due to COVID-19 pregnant women being at convalescence. However, a recent study examining placenta histological features from women acutely infected with SARS-CoV-2 at the time of birth found no significant differences in placental histopathologies, suggesting that COVID-19 does not directly affect inflammation at the maternal-fetal interface (Tasca et al., 2021).

Overall, our immune landscape data provide evidence that while the antibody and cellular components remain similar in pregnant and non-pregnant COVID-19 women during acute and convalescent phases of SARS-CoV-2 infection, perturbations of NK cell and unconventional T cell levels and inflammation are observed, providing key insights into further studies of immune responses in pregnancy. Our data will help inform patient management and education of COVID-19 during pregnancy.

## Data Availability

The source data underlying Figures and Supplementary Figures will be provided as Source Data. Raw FACS data are shown in the manuscript. FACS-source files are available from the authors upon request.

## ACKNOWLEDGMENTS

We thank all the participants involved in the study; Jeni Mitchell, Zelda Williams, Veronica Link, Beverly Cox, Robyn Esterbauer, Hannah Kelly, Jane Batten and Helen Kent for support with the cohort; Jill Garlick, Janine Roney, Anne Paterson and the research nurses at the Alfred Hospital. We acknowledge all DRASTIC (The use of cytokines as a preDictoR of disease Severity in criTically Ill Covid-patients) investigators from Austin Health, and thank the participants involved. The authors thank Effie Mouhtouris and Ana Copaescu for laboratory work and study coordination for the DRASTIC study, and George Drewett for patient recruitment. This research included samples and data from the Sentinel Travelers Research Preparedness Platform for Emerging Infectious Diseases (SETREP-ID). We acknowledge all SETREP-ID investigators and sites, and thank all participants involved. The authors thank Barbara Scher for setting up the ethics and governance for the SETREP-ID platform and the Australian Partnership for Preparedness Research for Infectious Disease Emergencies (APPRISE) for ongoing funding of SETREP-ID, Ajantha Rhodes, Judy Chang, Ashanti Dantanarayana and Rosalyn Cao who contributed to the SETREP-ID biobank. SETREP-ID is supported by funding through the National Health and Medical Research Council Centre of Research Excellence (NHMRC CRE), the Australian Partnership for Preparedness Research on Infectious Disease Emergencies (APPRISE AppID 1116530), the Snow Medical Foundation, the Jack Ma Foundation and the A2 Milk Company. We thank Leo Lee and Francesca Mordant for their assistance with the MN assays. This work was supported by the Australian National Health and Medical Research Council (NHMRC) Leadership Investigator Grant to KK (#1173871), Research Grants Council of the Hong Kong Special Administrative Region, China (#T11-712/19-N) to KK, the Jack Ma Foundation to KK, KS, DIG, IT and AWC, the Victorian Government MRFF award (#2002073) to SJK, DIG and AWC, MRFF Award (#1202445) to KK, MRFF Award (#2005544) to KK, SJK, AWC, ACC, DW and JAJ, NHMRC program grant 1149990 (SJK), NHMRC Program Grant (1113293) to DIG. THON was supported by a NHMRC Emerging Leadership Level 1 Investigator Grant (#1194036), HFK, CLG and JAT by NHMRC Early Career Fellowships (#1160333, #1160963 and #1139902), KS by a NHMRC Investigator grant (#1177174), DIG by a NHMRC Senior Principal Research Fellowship (#1117766), AWC by an NHMRC Career Development Fellowship (#1140509) and SJK by NHMRC Senior Principal Research Fellowship (#1136322). JRH, LH and WZ are supported by the Melbourne International Research Scholarship (MIRS) and the Melbourne International Fee Remission Scholarship (MIFRS) from The University of Melbourne. XJ was supported by China Scholarship Council-University of Melbourne joint Scholarship. JAJ is supported by an NHMRC Early Career Fellowship (ECF) (#1123673). KK and AWC were supported by the University of Melbourne Dame Kate Campbell Fellowship. The Melbourne WHO Collaborating Centre for Reference and Research on Influenza is supported by the Australian Government Department of Health.

## AUTHOR CONTRIBUTIONS

KK and LCR supervised the study. KK, LCR, JRH, BYC, LK, HK, SN and AWC designed the experiments. JRH, BYC, LK, KJS, TD, ERH, THON, HK, SN, XJ, LFA, LH, WZ, CES, JAN, HT and LCR performed and analysed experiments. TD, LH and HM analysed data. FA, FK and AKW provided reagents. KW, JAJ, AKW, GP, JA, IT, JD, ACC, SYCT, KB, DAW, FJ, NEH, OCS, JAT, CLG, CLW, SJK and ML recruited the patient cohorts and provided clinical data. JRH, SN, KS, DIG, AWC, SJK, ML, LCR and KK provided intellectual input into the study design and data interpretation. JRH, LCR and KK wrote the manuscript. All authors reviewed and approved the manuscript.

## METHODS

### Study participants and ethics statement

86 subjects were recruited into this study (Supp Table 1 and 2). Pregnant and non-pregnant women with acute or convalescent COVID-19 were recruited via the Mercy Hospital for Women, Royal Women’s Hospital, Royal Melbourne Hospital, Austin Hospital, Alfred Hospital or The University of Melbourne. Healthy pregnant donors were recruited via the Mercy Hospital for Women and the University of Melbourne. Healthy non-pregnant donors were recruited via The University of Melbourne or buffy packs obtained from the Australian Red Cross LifeBlood (West Melbourne, Australia). Peripheral or cord blood was collected in heparinized tubes and peripheral blood monocular cells (PBMCs) were isolated via Ficoll-Paque separation. Plasma was obtained from whole blood by centrifugation of heparin blood tubes at 300*g* for 10 min. Placenta samples were obtained and processed into single-cell suspension essentially as described with minor modifications(Koutsakos et al., 2018). Briefly, mononuclear cells were isolated and cryopreserved from placentae by mechanical dissociation and enzymatic digestion with 2mg/ml of Collagenase D (Roche) in RPMI containing 0.2 mg/mL DNase I (Roche), 1 mM HEPES, penicillin and streptomycin for 1 hour at 37°C. Cells were filtered through a 70µm strainer and red blood cells were lysed using a solution of 0.168M NH_4_Cl, 0.01mM EDTA and 12mM NaHCO_3_ in ddH_2_O. This study was a part of a larger study to understand immune responses to COVID-19 and immune perturbation during severe COVID-19.

Experiments conformed to the Declaration of Helsinki Principles and the Australian National Health and Medical Research Council Code of Practice. Written informed consents were obtained from all blood donors prior to the study. The study was approved by the Alfred Hospital (#280/14), Melbourne Health (HREC/66341/MH-2020 and HREC/17/MH/53), Austin Health (HREC/63201/Austin-2020), Monash Health (HREC/15/MonH/64), Mercy Health (R14/25 and R04/29), Australian Red Cross Lifeblood (2015#08), and the University of Melbourne (#1442952, #1749349, #2056901, #1443540, #2056761, #1955465, 2020-20782-12450-1, 2021-13973-14410-3 and 2021-13973-14410-3) Human Research Ethics Committees.

### Flow cytometry of whole blood, PBMCs and placenta

Fresh whole blood, PBMCs isolated from whole blood or placenta single-cell suspensions were used to assess cellular immunity, as previously described (Thevarajan et al., 2020). Four antibody panels were used to determine activation of (1) monocytes, T, B, NK and γδ T cells, (2) T_FH_ and ASC cell activation, (3) cytotoxicity profiles of T cells and NK cells expressing intracellular granzymes A, B, K and M and perforin, and (4) activation and phenotypes of MAIT and γδ T cells (Supp Fig 6 and 7). Panels 1 and 2 were previously described (Koutsakos et al., 2021); panel 3 and 4 in Supp Table 4. Cells were stained, RBC lysed if from whole blood, then fixed in 1% PFA, or stained intracellularly using the eBioscience™ Foxp3/Transcription Factor Staining Buffer Set (Thermo Fisher Scientific, Carlsbad, CA, USA), as previously described (Thevarajan et al., 2020). Samples were acquired on a LSRII Fortessa (BD Biosciences) and analyzed using FlowJo v10 software.

### SARS-CoV-2 Receptor Binding Domain and Nucleocapsid ELISA

ELISA for the detection of RBD- or N-specific IgG, IgM and IgA antibodies were performed as previously described (Amanat et al., 2020; Koutsakos et al., 2021; Rowntree et al., 2021), using flat bottom Nunc MaxiSorp 96-well plates (Thermo Fisher Scientific) for antigen coating (2µg/ml), blocking with PBS (with w/v 1% BSA) and serial dilutions in PBS (with v/v 0.05% Tween and w/v 0.5% BSA). Endpoint titres were determined by interpolation from a sigmoidal curve fit (all R-squared values >0.95; GraphPad Prism 9) as the reciprocal dilution of plasma that produced >15% (for IgA and IgG) or >30% (for IgM) absorbance of the positive control at a 1:31.6 (IgG and IgM) or 1:10 dilution (IgA). Seroconversion was defined as any titre greater than the mean plus two standard deviations of non-COVID-19 control plasma samples.

### Antibody avidity ELISA

The avidity of RBD-specific IgG and IgM were assessed by urea-mediated dissociation ELISA. Nunc Immuno MaxiSorp flat-bottom 96-well plates (Thermo Fisher Scientific) were coated with RBD protein overnight at 4°C. Plates were washed and blocked with PBS (with w/v 1% BSA) for at least 1 h. Donor plasma was added in log_0.5_ dilutions and incubated for 2 h at room temperature. Wells were washed and 6M urea added and incubated for 15 min. Bound antibodies were then detected using either HRP-conjugated anti-human IgG or anti-human IgM antibody as described previously. The amount (in percentage) of antibody remaining was determined by comparing the total area of the antibody titration curve (across 4 dilutions) in the presence and absence of urea treatment and is expressed as the avidity index.

### Surrogate virus neutralization test

Surrogate virus neutralization test ELISA (GenScript, NJ, USA) for the detection of antibodies that block the interaction between the SARS-CoV-2 spike protein RBD and the host receptor ACE2 was carried out as previously described (Rowntree et al., 2021). HRP-conjugated recombinant SARS-CoV-2 RBD fragment bound to any circulating neutralizing antibodies to RBD preventing capture by the human ACE2 protein in the well, which was subsequently removed in the following wash step. Substrate reaction incubation time was 20 mins at room temperature and results were read by spectrophotometry. Colour intensity was inversely dependent on the titre of anti-SARS-CoV-2 neutralizing antibodies.

### Microneutralization test

Microneutralization activity of plasma samples was determined essentially as described (Juno et al., 2020a). Vero cells were used for the propagation of the SARS-CoV-2 isolate CoV/Australia/VIC01/2020 (Caly et al., 2020), stored at -80°C. Heat inactivated sera (56°C for 30 min) was serially diluted and serum/virus mixtures assessed for residual virus infectivity in quadruplicate wells of Vero cells incubated in serum-free media containing 1μg/ml of TPCK trypsin at 37°C and 5% CO_2_. Viral cytopathic effect was read on day 5. The neutralizing antibody titer was calculated using the Reed–Muench method, as described (Juno et al., 2020a).

### Total IgG glycosylation

Total IgG glycosylation was analysed as previously described using capillary electrophoresis (Mahan et al., 2015). Briefly, Melon gel IgG purification resin was used to purify total IgG from plasma according to the manufacturer’s protocol (Thermo Fisher). N-linked glycans on purified IgG was analysed using LabChip GXII Touch Microchip-CE platform per manufacturer’s protocol (Perkin Elmer).

### Coupling of carboxylated beads

A custom multiplex bead array was designed and coupled with SARS-CoV-1 and SARS-CoV-2 spike 1 (stem, Sino Biological), spike 2 (head, ACRO Biosystems), RBD (BEI Resources) and nucleoprotein (ACRO Biosystems) as previously described (Selva et al., 2021) (Supp Table 3). In addition, SARS-CoV-2 spike trimers (kindly provided by Adam K. Wheatley) and SARS-CoV-2 spike trimers (BPS Bioscience) were also included. Tetanus toxoid (Sigma-Aldrich), influenza hemagglutinin (H1Cal2009; Sino Biological) and SIV gp120 (Sino Biological) were included as positive and negative control antigens, respectively. Antigens were covalently coupled to magnetic carboxylated beads (Bio Rad) using a two-step carbodiimide reaction and blocked with 0.1% BSA, before being resuspended and stored in PBS 0.05% sodium azide until use.

### Luminex bead-based multiplex assay

A custom multiplex assay was used to investigate the isotypes and subclasses of SARS-CoV-1 and -2 specific antibodies present in plasma samples (Selva et al., 2021). In brief, 20µl of working bead mixture (1000 beads per bead region) and 20µl of diluted plasma (final dilution 1:200) were added per well and incubated overnight at 4°C on a shaker. Fourteen different detectors were used to assess pathogen-specific antibodies. Single-step detection was done using phycoerythrin (PE)-conjugated mouse anti-human pan-IgG, IgG1-4 and IgA1-2 (Southern Biotech; 1.3µg/ml, 25µl/well). C1q protein (MP Biomedicals) was first biotinylated (Thermo Fisher Scientific), then tetramerized with Streptavidin R-PE (SA-PE; Thermo Fisher Scientific) before dimers were used for single-step detection. For the detection of FcγR-binding, soluble recombinant FcγR dimers which come in higher affinity (FcγRIIa-H131 and FcγRIIIa-V158) or lower affinity (FcγRIIa-R131, FcγRIIb and FcγRIIIa-F158; 1.3µg/ml, 25µl/well; kind gifts from Bruce D. Wines and P. Mark Hogarth) were first added to the beads, washed, and followed by the addition of SA-PE. For the detection of IgM, biotinylated mouse anti-human IgM (mab MT22; Mabtech; 1.3µg/ml, 25µl/well) was first added to beads, washed, followed by SA-PE. Assays were performed in duplicates and read on the Flexmap 3D.

### Data normalization for multiplex analysis

Tetanus, H1Cal2009, BSA and SIV control antigens were removed from the analysis. Low signal features were removed when the 75^th^ percentile response for the feature was lower than the 75^th^ percentile of the BSA positive control. Right shifting was performed on each feature (detector–antigen pair) individually if it contained any negative values, by adding the minimum value for that feature back to all samples within that feature. Right-shifted data were log-transformed using the following equation, where x is the right-shifted data and y is the right-shifted log-transformed data: y = log10(x + 1) to achieve normal distribution. Data were furthered normalized by mean centering and variance scaling each feature using the z-score function in Matlab in the subsequent multivariate analyses.

### Multivariable methods for identification of the key antibody features

A Least Absolute Shrinkage and Selection Operator (LASSO) penalised logistic regression model was used to determine the minimal set of features needed to predict pregnancy status during acute and convalescent COVID-19 (O’Brien, 2016). The LASSO shrinks data toward a simple, sparse model with fewer parameters and identifies the subset of antibody features that best discriminate between two groups. The frequency of selected features in resampling was considered as the criterion of variable importance (O’Brien, 2016). The feature selection stability was defined as the proportion of times that a feature was picked in the selected set of important features, when the model was repeatedly fitted to 1000 resampled subsets of data. Inner cross validations (CV) ranging from 4-fold to 10-fold were performed for each of the resampled datasets. Following model prediction, 10-fold CV was selected due to its consistency.

### Principal component analysis

PCA was performed in the Eigenvectors PLS toolbox (Eigenvector Research, Inc., Manson, USA) in MATLAB. PCA is an unsupervised technique that was used to visualise the variance in the samples based on all of the measured features and to reduce the dimensionality of the dataset (Jolliffe, 1986). Every Ab feature is assigned a loading, which in linear combinations creates a principal component (PC). Loadings and PCs are calculated to describe the maximum amount of variance in the dataset. Two-dimensional score plots were generated to visually assess separation between groups using their individual response measurements expressed through the PCs. The percent of variance described by each PC is a measure of the amount of variance in antibody response explained by that respective PC (Selva et al., 2021).

### Cytokine measurements

Plasma levels of IL-1β, IFN-α2, IFNγ, TNFα, MCP-1 (CCL2), IL-6, IL-8 (CXCL8), IL-10, IL-12p70, IL-17A, IL-18, IL-23 and IL-33 measured using the LEGENDplex™ Human Inflammation Panel 1 kit (BioLegend, San Diego, CA, USA). Donor plasma was diluted 1:2 and the assay was performed according to manufacturer’s instructions. Samples were acquired on a FACSCanto™ II cytometer (BD Biosciences) and analysed with the online QOGNIT LEGENDplex™ program.

### Statistical analysis

Data and statistical analysis were performed in GraphPad Prism (version 9). Non-linear regression plots were made in R studio (version 4) using the ggplot package (Wickham, 2016) and LOESS model. PESTLE and SPICE software (version 6.1) were used for analysis of cytotoxic granzymes and perforin in NK cell and T cell subsets (Roederer et al., 2011).

## FIGURE LEGENDS

**Supplementary Figure 1.**
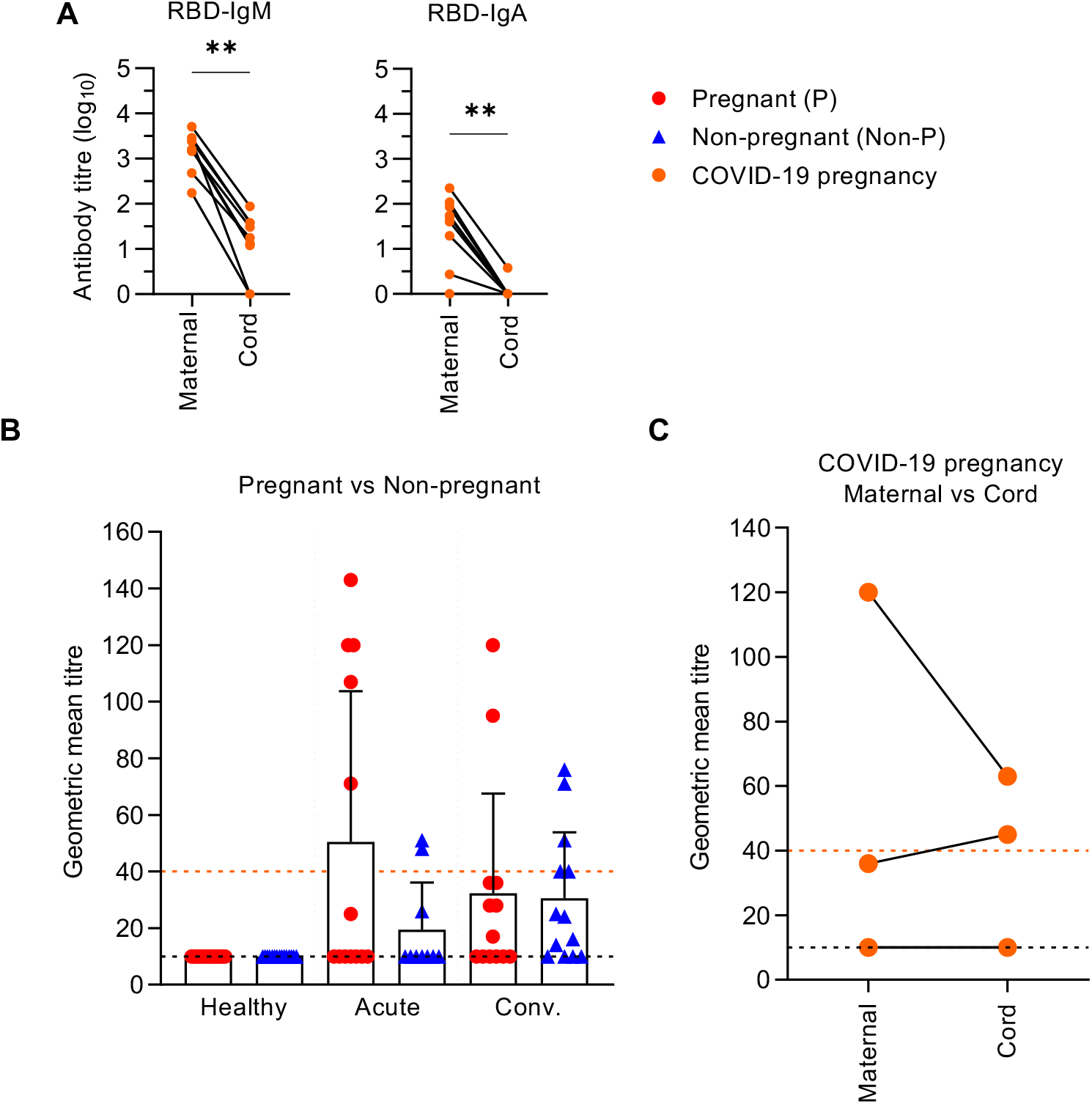

**Supplementary Figure 2.**
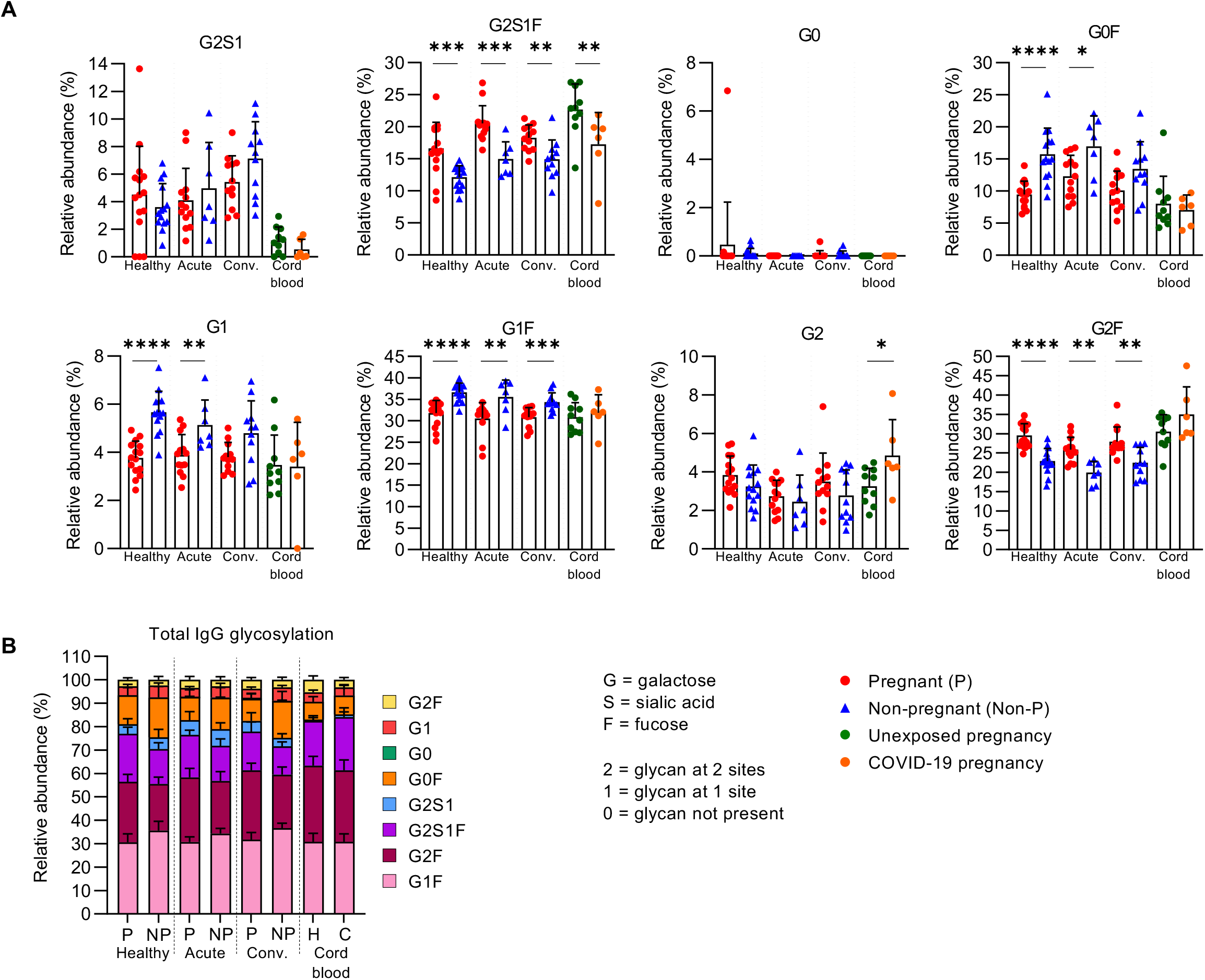

**Supplementary Figure 3.**
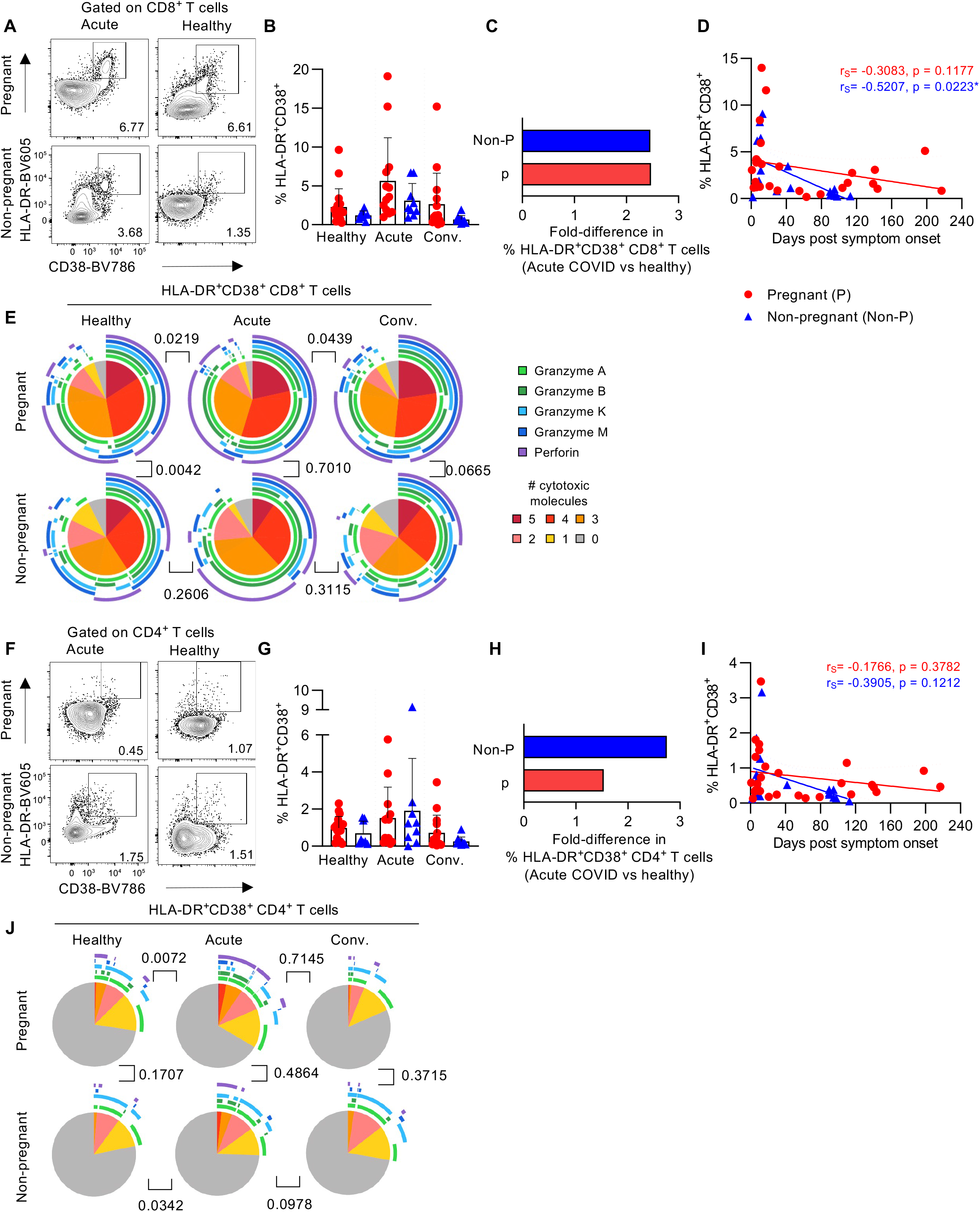

**Supplementary Figure 4.**
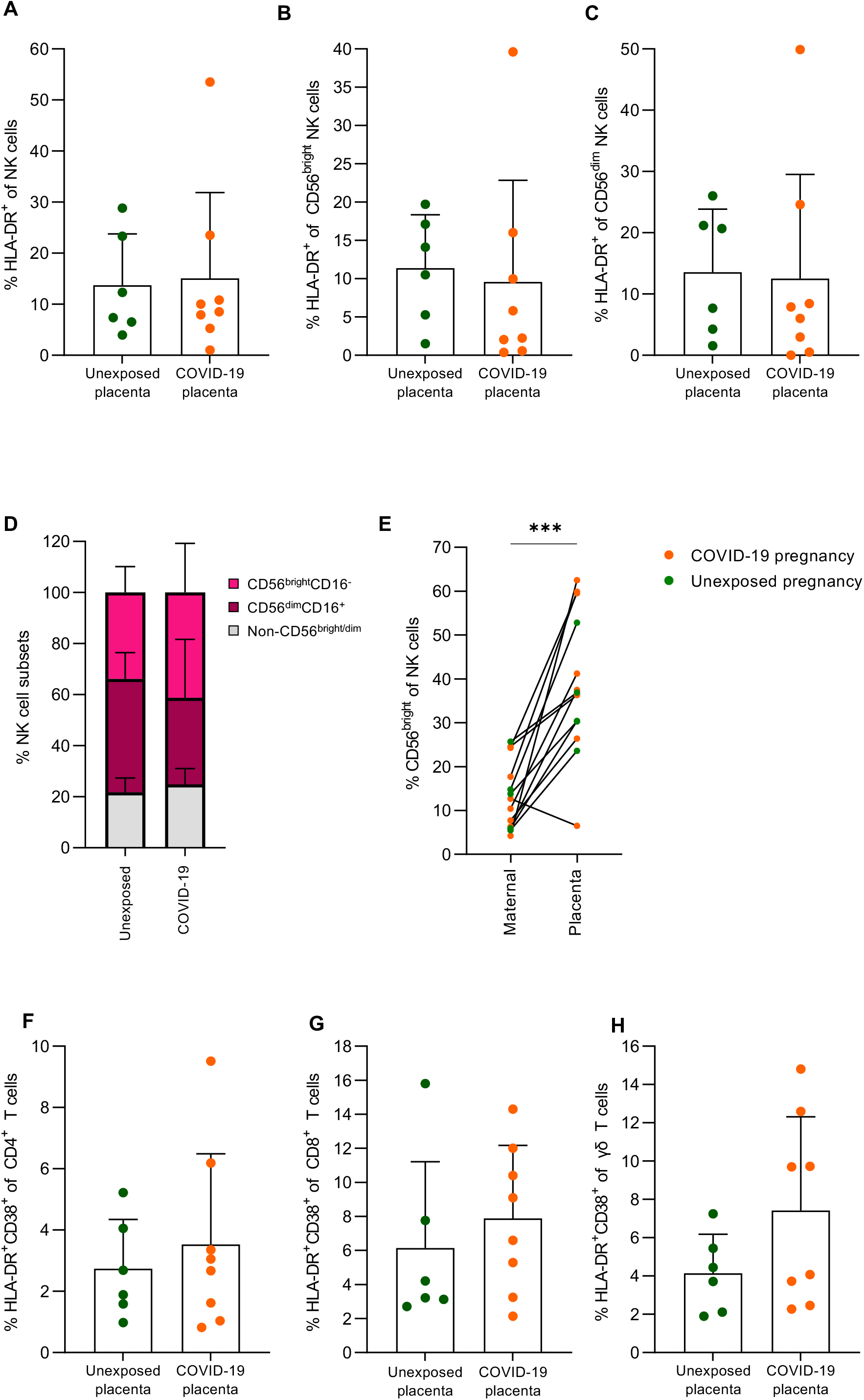

**Supplementary Figure 5.**
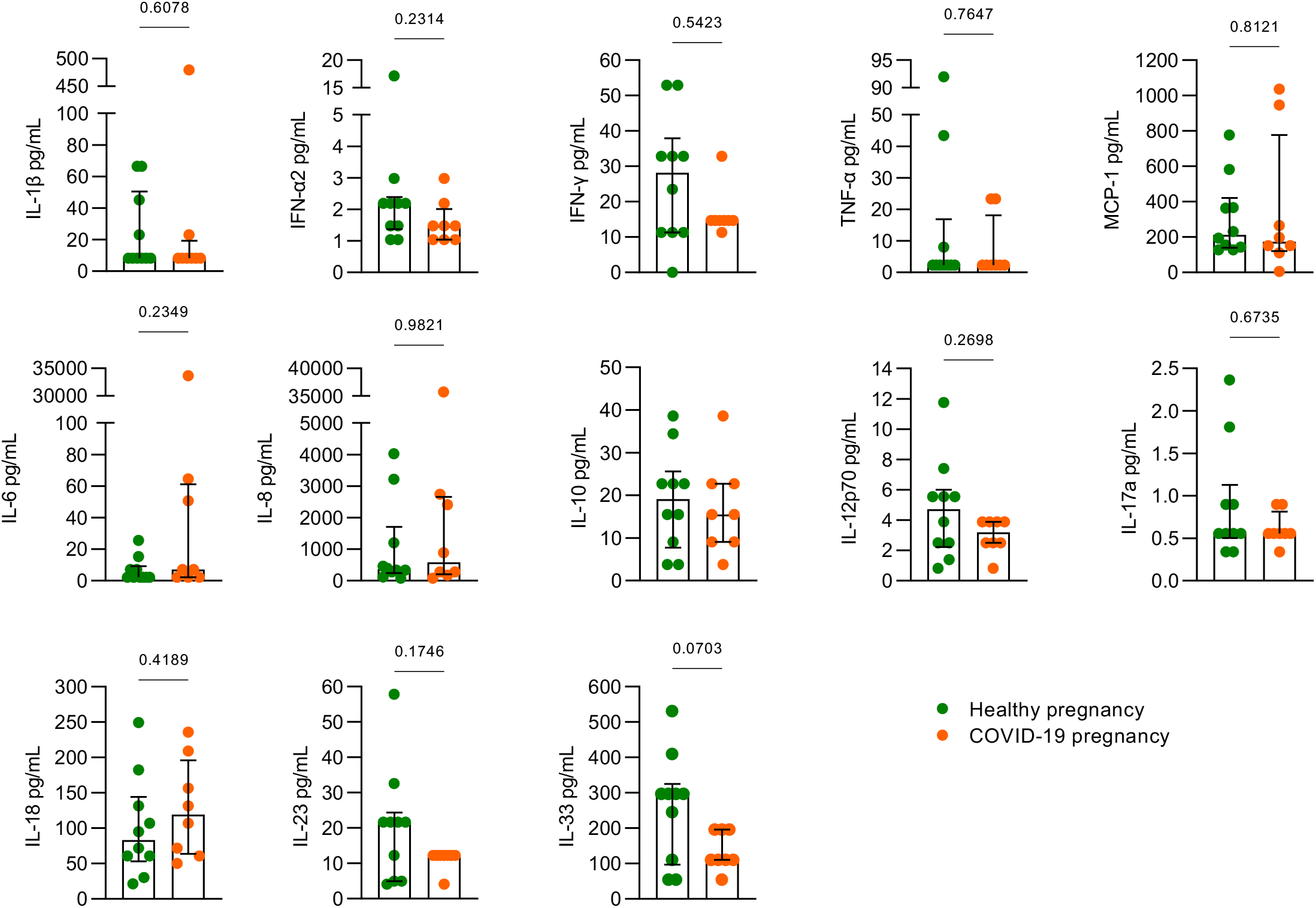

**Supplementary Figure 6.**
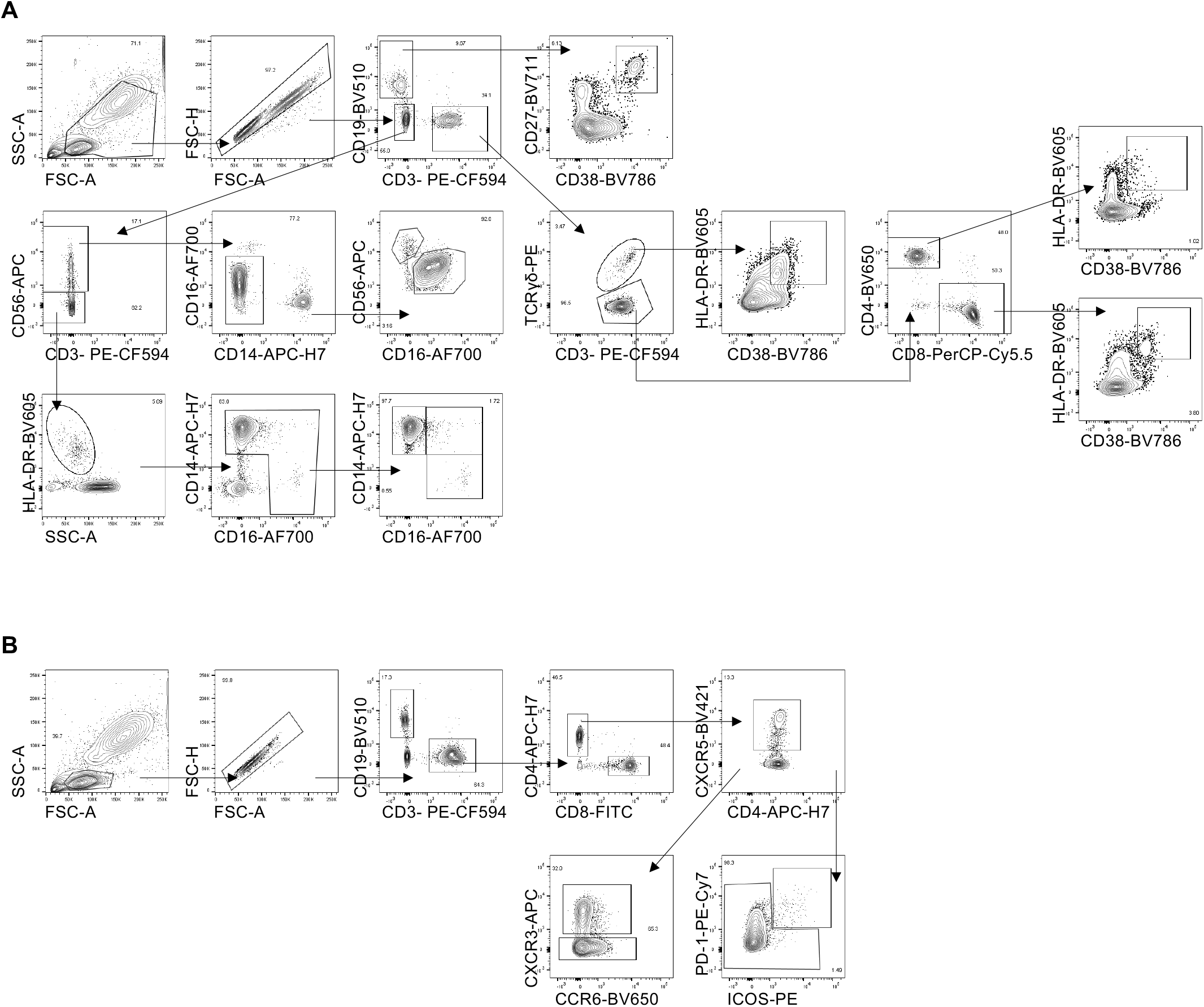

**Supplementary Figure 7.**
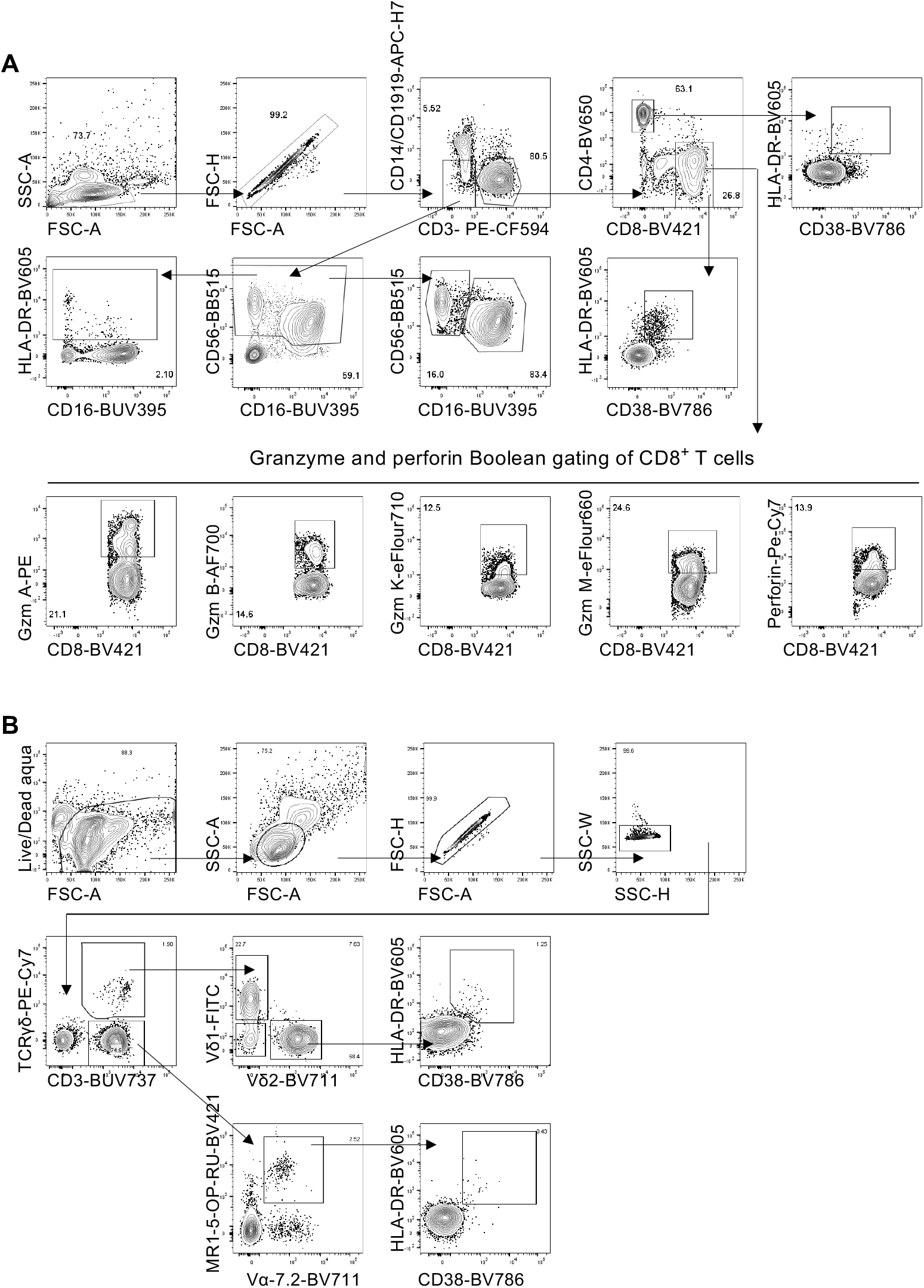

